# Constitutively active Lyn kinase causes a cutaneous small vessel vasculitis and liver fibrosis syndrome

**DOI:** 10.1101/2022.09.27.22280319

**Authors:** Adriana A. de Jesus, Guibin Chen, Dan Yang, Tomas Brdicka, Natasha Ruth, David Bennin, Dita Cebecauerova, Hana Malcova, Helen Freeman, Neil Martin, Karel Svojgr, Murray Paso, Farzana Bhuyan, Sara Alehashemi, Andre Rastegar, Kat Uss, Lela Kardova, Bernadette Marrero, Iris Duric, Ebun Omoyinmi, Petra Peldova, Chyi-Chia Richard Lee, David E. Kleiner, Colleen M. Hadigan, Stephen M. Hewitt, Stefania Pittaluga, Carmelo Carmona-Rivera, Katherine R. Calvo, Nirali Shah, Miroslava Balascakova, Danielle Fink, Radana Kotalova, Zuzana Parackova, Lucie Peterkova, Daniela Kuzilkova, Vit Campr, Lucie Sramkova, Angelique Biancotto, Stephen R. Brooks, Cameron Manes, Eric Meffre, Rebecca L. Harper, Hyesun Kuehn, Mariana J. Kaplan, Paul Brogan, Sergio D. Rosenzweig, Melinda Merchant, Zuoming Deng, Anna Huttenlocher, Susan Moir, Doug Kuhns, Manfred Boehm, Karolina Skvarova Kramarzova, Raphaela Goldbach-Mansky

## Abstract

Neutrophilic inflammation is a hallmark of many monogenic autoinflammatory diseases; pathomechanisms that regulate extravasation of damaging immune cells into surrounding tissues are poorly understood. We identified three unrelated boys with perinatal-onset of neutrophilic cutaneous small vessel vasculitis and systemic inflammation. Two patients developed liver fibrosis in their first year of life. Next-generation sequencing identified two *de novo* truncating variants in the Src-family tyrosine kinase, *LYN*, p.Y508*, p.Q507* and a *de novo* missense variant, p.Y508F, that result in constitutive activation of Lyn kinase. Functional studies reveal increased expression of ICAM-1 on induced patient-derived endothelial cells (iECs) and of β2-integrins on neutrophils that increase neutrophil adhesion and vascular transendothelial migration (TEM). Treatment with TNF inhibition improved systemic inflammation; and liver fibrosis resolved on treatment with the Src kinase inhibitor dasatinib. These findings reveal a critical role for Lyn kinase in modulating inflammatory signals, regulating microvascular permeability and neutrophil recruitment, and in promoting hepatic fibrosis.

## Summary

Next-generation sequencing (NGS) of disease probands and their parents has propelled the discovery of monogenic causes of autoinflammatory diseases,^1^ that led to the identification of key innate immune pathways as cause for severe clinical phenotypes, and generated novel targets for drug development. Here, we describe three unrelated boys with three *de novo* mutations in the gene encoding the Src-family tyrosine kinase, Lyn kinase, *LYN;* all presented with systemic inflammation and recurrent neutrophilic small vessel vasculitis. Two patients with truncating mutations had liver fibrosis, that resolved in one patient on treatment with the Src kinase inhibitor dasatinib.

We performed trio whole-exome sequencing (WES) on whole blood from Patient 1 and his parents, and identified a nonsense germline mutation in a coding region of the Src kinase, *LYN*, c.1524C>G, p.Y508* (transcript NM_002350), that resulted in truncation of five terminal amino acids, including a regulatory tyrosine at position p.Y508. In Patient 2, a targeted NGS panel revealed a missense mutation in *LYN* c.1523A>T, resulting in the replacement of tyrosine at position 508 by a phenylalanine (p.Y508F), and clinical WES in Patient 3 identified a nonsense germline mutation in *LYN*, c.1519C>T resulting in loss of six terminal amino acids including p.Y508. The three mutations occurred *de novo* (Fig. 1a,b and Supplementary Fig. 1), and were predicted to be deleterious. All were absent in public databases including gnomAD. *LYN* variant features, including amino acid conservation and pathogenicity predictions are detailed in Supplementary Table 1.

**Figure 1.**
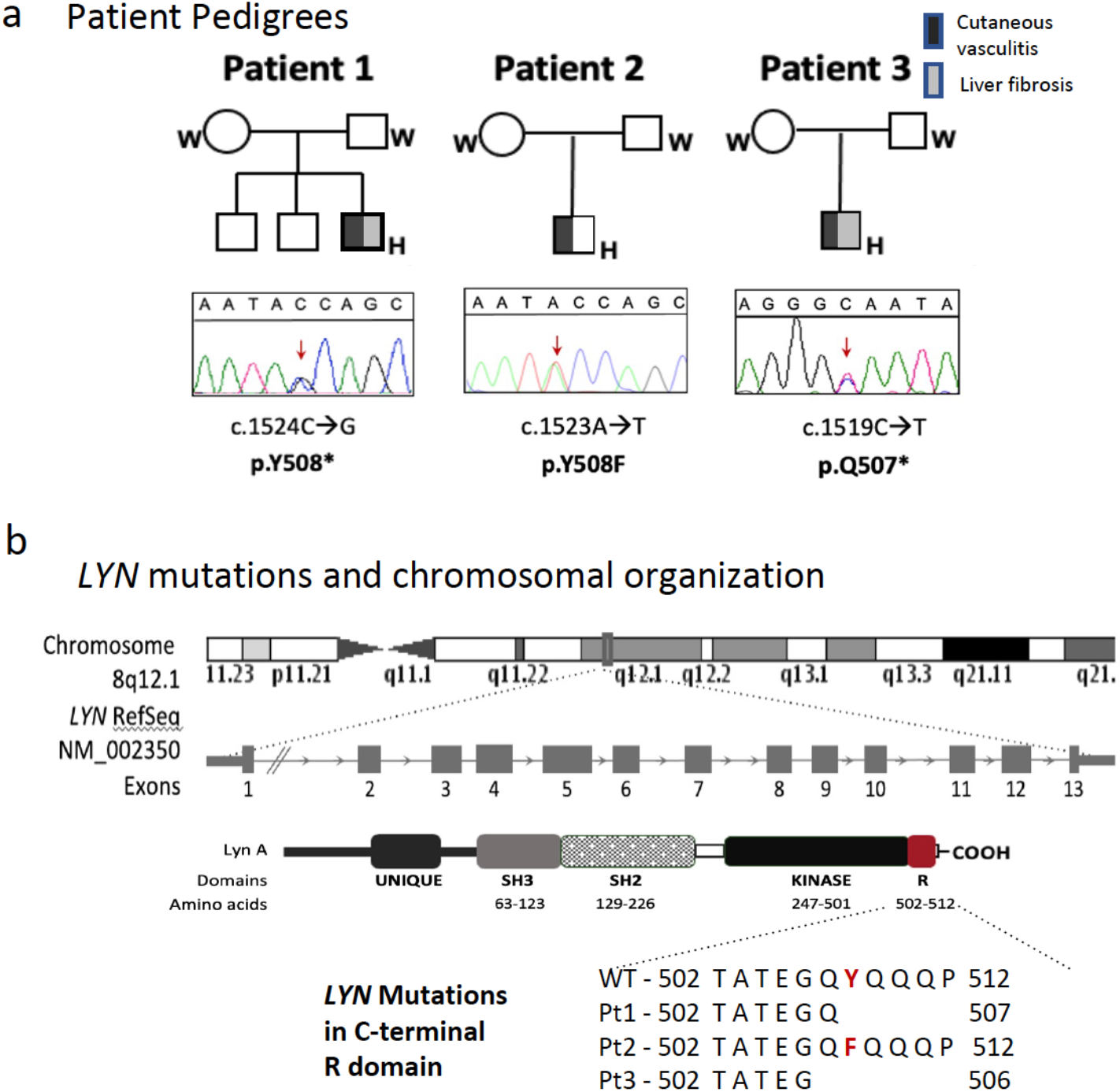
Gain-of-Function mutations in *LYN* leads to novel autoinflammatory syndrome. **a**. The pedigrees show a *de novo* mutation in *LYN* in each of the three patients. Squares and circles represent male and female family members, respectively; solid symbols and open symbols indicate affected and unaffected family members, respectively. **b**. In Patients 1 and 3 the truncating mutations p.Y508* and p.Q507* result in the loss of the 5 or 6 terminal amino acids, respectively; and in Patient 2, the amino acid substitution from a tyrosine to a phenylalanine prevents phosphorylation in the C-terminal regulatory domain of Lyn kinase.

All three patients developed diffuse purpuric skin lesions (Fig. 2a,b), fever, and increased C-reactive protein (CRP) in infancy. Patient 1 and Patient 3 had hepatosplenomegaly, transaminitis, and severe thrombocytopenia. Patient 1 underwent a splenectomy for thrombocytopenia and developed leukocytosis, thrombocytosis and persistent anemia post-splenectomy. He also had transiently elevated circulating autoantibodies including ANA, anti-Sm, anti-SSA, anti-mitochondrial and anti-phospholipid antibodies without clinical autoimmune manifestations; antibody titers became negative after a course of corticosteroids. Persistently elevated liver function tests (LFTs) led to a liver biopsy, which showed a mild periportal lymphocytic infiltrate, clusters of macrophages and scattered neutrophils within hepatic sinuses and portal areas, marked biliary ductopenia and early peri-sinusoidal fibrosis (Fig.2c). Other clinical manifestations included intermittent abdominal and testicular pain, headaches, conjunctival and periorbital erythema, arthralgias, and myalgias worse after exercise or trauma. A magnetic resonance imaging (MRI) during myalgia showed fasciitis and muscle edema (Fig. 2a). Patient 3 had intrauterine growth restriction (IUGR) and congenital hydrocele. Liver elastography was consistent with liver cirrhosis. His thrombocytopenia improved on etanercept and initiation of dasatinib is considered if transaminitis and high elastography scores do not improve. More clinical and laboratory features of the three patients can be provided to the readers upon request.

**Figure 2.**
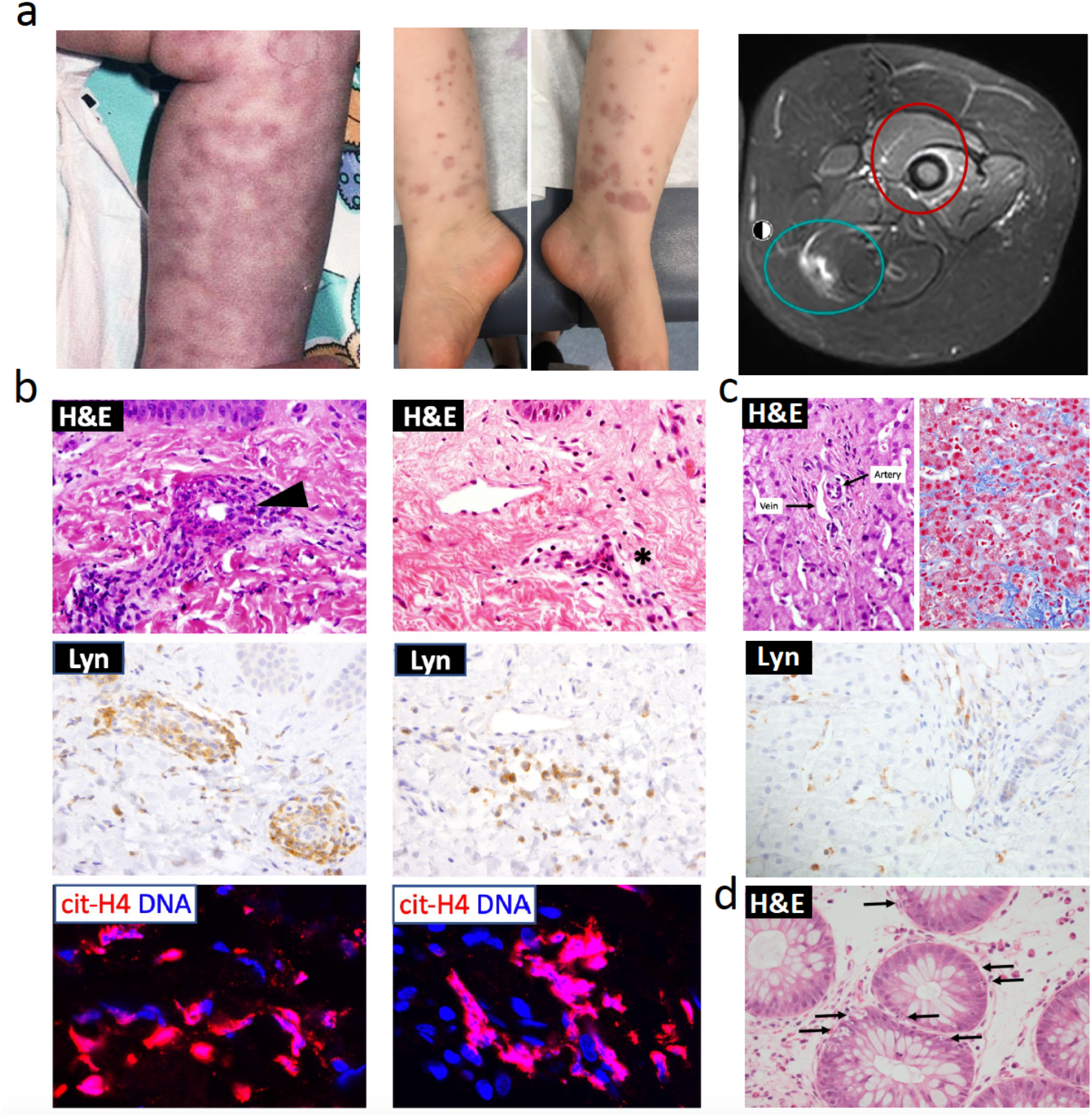
Clinical and histopathologic features of patients with *LYN* gain-of-function mutations. **a**. Non-blanching vascular rashes in Patient 2’s (left panel) and Patient 1’s (center panel) lower extremities, and left thigh MRI from Patient 1 (right panel) depicting fasciitis (green circle) and periostitis (red circle) are shown. **b**. Hematoxylin & eosin (H&E) staining from lesional skin biopsies of Patients 1 and 2 show destruction of a small vessel with surrounding inflammatory cells in Patient 1 (black arrow) and a perivascular infiltrate in patient 2 (asterisk) (upper panels). The endothelial cells and surrounding neutrophils are Lyn kinase (Lyn) positive in both patients (middle panels). The lower panels show the localized accumulation of NETs (pink) surrounding the small vessels in Patient 1 and 2 respectively. cit-H4, citrulline histone H4. **c**. A Liver biopsy performed in Patient illustrates a portal area lacking a bile duct on H&E (left upper panel) and early perisinusoidal fibrosis (right upper panel) on Masson’s trichrome staining (right upper panel). Lyn staining of sinusoidal cells and endothelial cells in the portal areas is shown in Patient 1’s liver biopsy (lower panel). **d**. Colon biopsy from Patient 2 shows apoptosis of multiple crypt epithelial cells (black arrows).

The clinical presentation of Lyn gain-of-function (GOF) mutations as autoinflammatory disease with sterile neutrophilic small vessel vasculitis was unexpected, as mice engineered to carry the p.Y508F mutation, that is disease-causing in Patient 2, develop high-titer autoantibodies and glomerulonephritis reminiscent of systemic lupus erythematosus.^2^ Although clinical tests have been negative, Patient 2 developed transiently elevated autoantibodies and in vitro B cell assays suggested defects in central and peripheral B cell tolerance (see Supplementary Results). All patients had histopathological evaluation of lesional skin biopsies, which showed a dense neutrophilic infiltrate around small vessels including capillaries (Fig. 2a, b and Supplementary Fig. 2) suggestive of pauci-immune small vessel vasculitis (ANCA-negative). Extravasated neutrophils form neutrophil extracellular traps (NETs) in areas of loss of vessel wall integrity (Fig. 2b and Supplementary Fig. 2). Lyn kinase is expressed in endothelial cells of small vessels, in neutrophils, monocytes and macrophages (Fig. 2a, Supplementary Fig. 2a,3) and in liver sinusoidal endothelial cells (LSECs) (Fig. 2c). Patient 1 and Patient 2 developed both post-infectious diarrhea that resolved without treatment; colon biopsies showed apoptotic crypt injury, reminiscent of graft-versus-host disease (Fig. 2d).

To further investigate the functional impact of the *de novo* mutations, we assessed Lyn kinase phosphorylation. The C-terminal tail tyrosine residue, p.Y508, inactivates wildtype Lyn kinase by association with its own SH2 domain (Fig. 3a).^3,4^ We hypothesized that the C-terminal deletions of Lyn kinase Y508, or the Y508F substitution increase Lyn kinase activity (Fig. 3a, left panel).^5^ Transfection of wildtype and mutant Lyn into HEK293FT cells showed constitutive phosphorylation of the kinase activating tyrosine, p.Y397, and absent phosphorylation of the “inhibitory tyrosine”, p.Y508 (Fig. 3a). Mutant Lyn kinase caused increased phosphorylation of several Lyn kinase targets (Fig. 3b). Flow cytometry analysis of anti-IgM stimulated B cells from Patient 1 showed increased phosphorylation of Lyn kinase substrates, including PLCγ2, CD19, CD79A that was blocked by the Src kinase inhibitors dasatinib and PP2 (Supplementary Fig. 4). Serologic analysis from untreated Patients 1, 2 and 3 showed elevated pro-inflammatory cytokines (i.e. IL-6), and/or markers of neutrophil (i.e. L-selectin and lipocalin), and endothelial cell activation (i.e. ICAM-1 and E-selectin) (Fig. 3c). Monocyte-derived macrophages, upon LPS stimulation, released increased levels of cytokines TNF-α and IL-6, and chemokines CXCL10 and CCL3/4/5, and shed increased levels of sICAM-1 when compared to healthy and disease controls (Supplementary Fig. 5). Neutrophils showed signs of constitutive activation, including low surface expression of CD62L and high expression of β2-integrin adhesion molecules, with normal cytokine responses to a broad range of endogenous, Toll-like receptor (TLR) and microbial stimulants (Supplementary Fig. 6 and Supplementary Table 2). In lesional skin biopsies, ICAM-1 was upregulated and colocalized with the endothelial marker CD31/PECAM along the endothelial wall (Supplementary Fig. 2b).

**Fig. 3.**
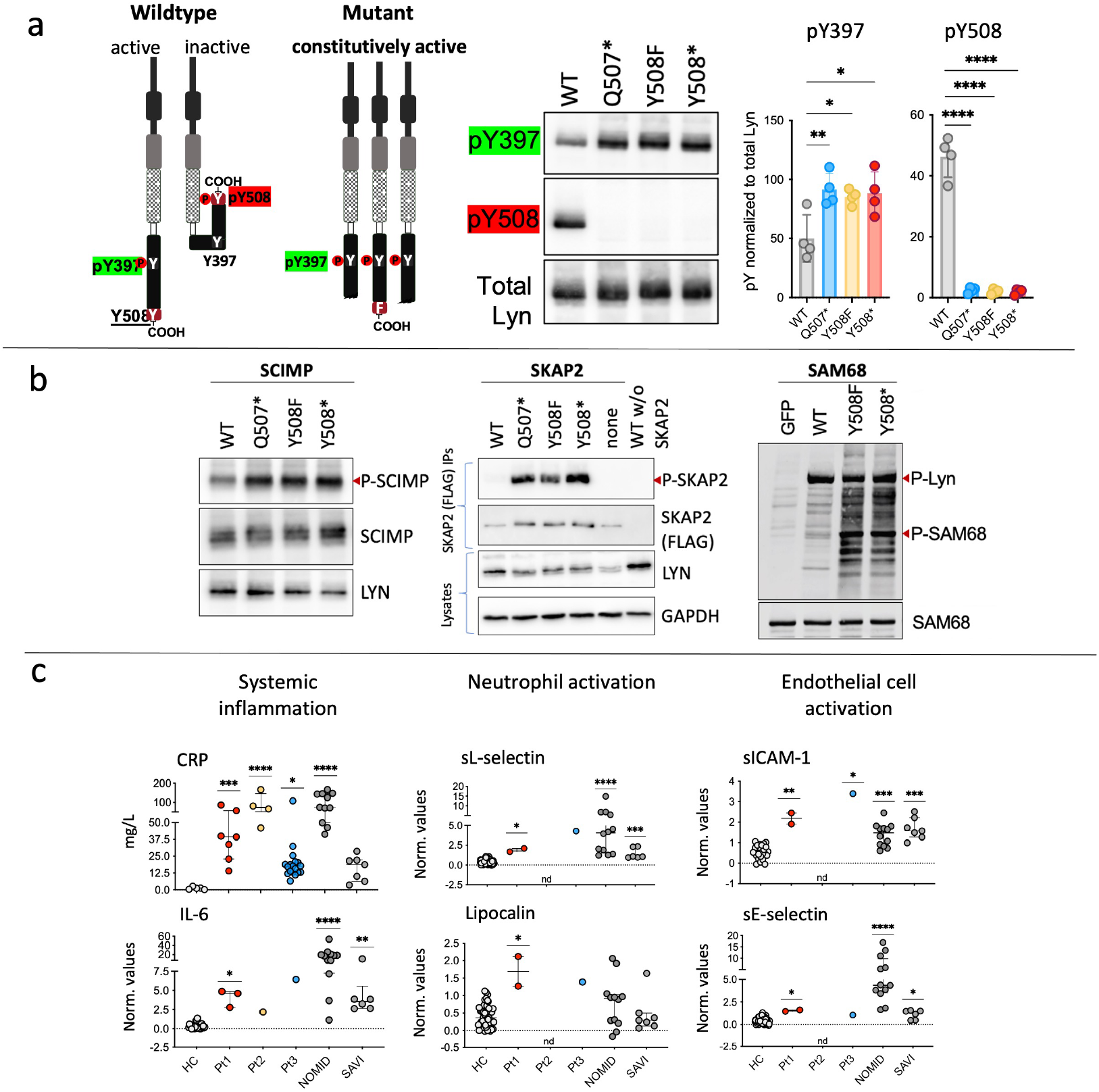
Constitutive Lyn kinase activation and evidence of systemic inflammation and endothelial activation. **a**. Active and inactive configuration of Lyn kinase depends on phosphorylation of an inhibitory tyrosine (Y) at position Y508. The *LYN* mutations detected in Patients 1, 2 and 3 lead to Lyn constitutive activity (left panel). Western blot of lysates from HEK293FT cells that were transiently transfected with wildtype (WT) or mutant *LYN* haboring c.1524C>G (p.Q507*), c.1523A>T (p.Y508F), or c.1519C>T (p.Y508) variants respectively, demonstrate increased phosphorylation of p.Y397 and absent phosphorylation of p.Y508 in the mutant constructs (right panel). * p < 0.05, ** p < 0.01, **** p < 0.0001 as determined by One Way ANOVA with Bonferroni’s Multiple Comparison Post-test. **b**. To assess Lyn phosphorylation of the adaptor proteins, Scimp, Skap2 and SAM68, HEK293FT cells were transiently co-transfected with wildtype (WT) or mutant *LYN* and with *SCIMP* or FLAG-tagged *SKAP2* (left and center panels). PLB-985 cells were stably transfected with an empty pMX-Lyn-GFP construct containing a GFP tag (GFP), or constructs containing wildtype (WT), or mutant *LYN* (right panel). Red arrows indicate phosphorylated Lyn kinase or phosphorylated substrates of Lyn kinase. **c**. Patients had increased CRP and IL-6 levels, comparable to patients with other systemic autoinflammatory diseases SAIDs, NOMID and SAVI (left panel). Patients 1 and 3 had marked elevation of biomarkers of neutrophil (sL-selectin and lipocalin) (center panel) and endothelial cell (sICAM-1 and sE-selectin) (right panel) activation, comparable to patients with NOMID and SAVI. NOMID, neonatal-onset multisystem inflammatory disorder; SAVI, STING-associated vasculopathy with onset in infancy. * p < 0.05, ** p < 0.01, *** p<0.001, **** p < 0.0001 as determined by Kruskal-Wallis test.

All patients responded partially (Patient 1 and Patient 3) or fully (Patient 2) to treatment with the TNF inhibitor etanercept after failing treatment with glucocorticosteroids and intravenous immunoglobulin (IVIG) (Patient 1), or IL-1 and IL-6 targeted treatment (Patient 2). Patient 2 had partial responses to colchicine but recurrence of rash, fatigue and systemic inflammation led to empirical treatment with the TNF inhibitor etanercept, which rapidly improved the skin rashes and normalized the acute phase reactants and led to long-term remission (Fig. 4a). Patient 1 initially received glucocorticosteroids and IVIG, but after 6 months on treatment, he continued to experience painful rashes, and fatigue and a repeat liver biopsy showed progressive ductopenia involving 50% of portal areas and development of bridging fibrosis (Fig. 4b, Supplementary Table 3). The discovery of the GOF mutation in *LYN* and the disease progression prompted treatment with the Src kinase inhibitor dasatinib, which blocks Lyn kinase, and gradually normalized CRP (Fig.4a), and LFTs (Fig. 4b), as well as inflammatory cytokines and chemokines in peripheral blood (Fig. 4c). Temporary discontinuation of dasatinib resulted in skin rashes with a rise in inflammatory markers and in LFTs, that resolved on reinstitution of dasatinib (Fig. 4a-c). After 30 months on dasatinib monotherapy, bile ducts were reconstituted, and liver fibrosis had regressed (Fig. 4b, d and Supplementary Fig. S7). The patient has been in inflammatory remission over the last 3.5 years. Patient 3, was started on etanercept and had partial improvement of systemic inflammation, thrombocytopenia and direct hyperbilirubinemia but persistently elevated LFTs (Fig. 2b).

**Figure 4.**
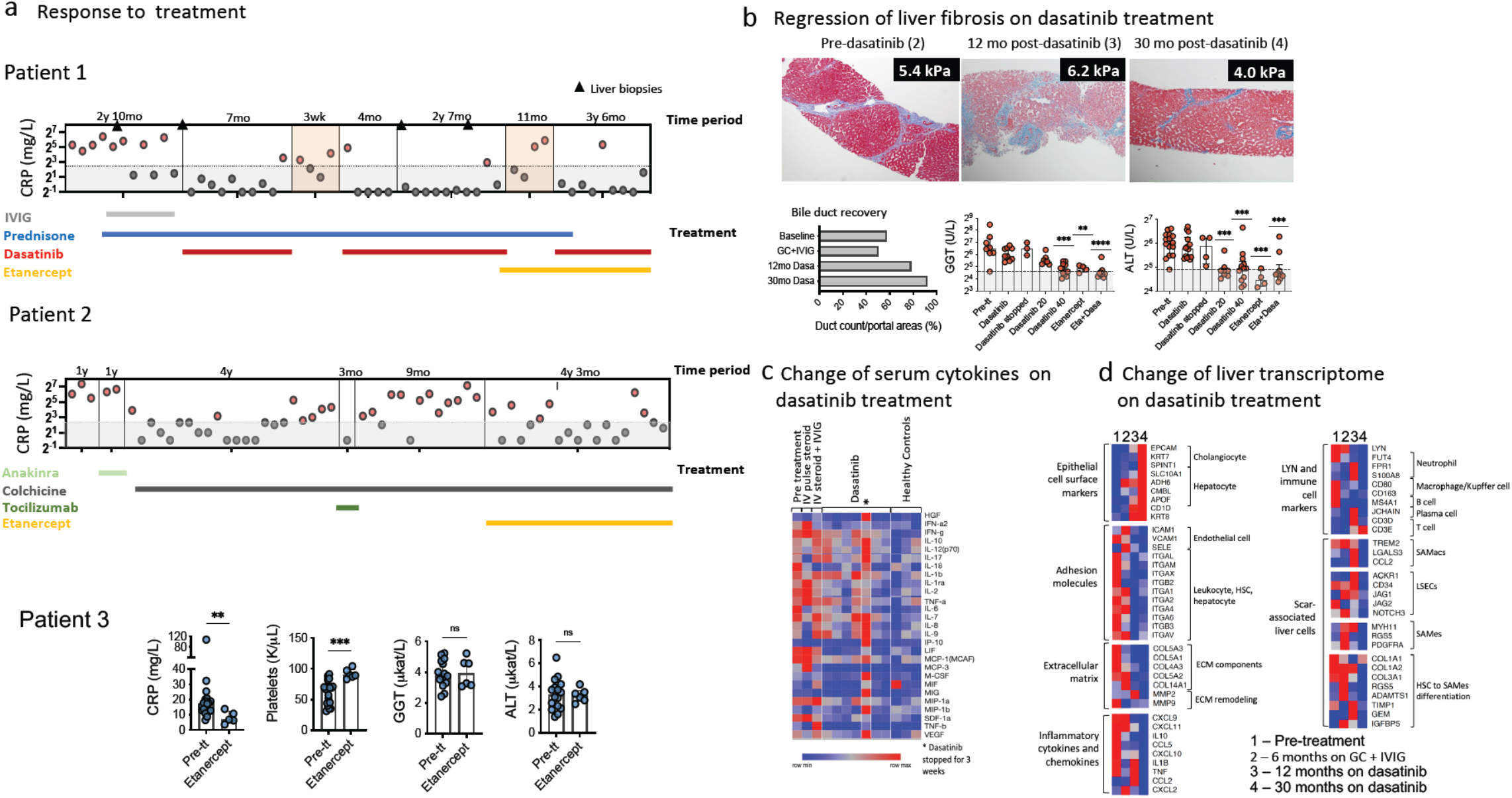
Response to treatment in the three patients with gain-of-function mutations in *LYN*. **a**. Longitudinally obtained CRP levels are depicted for all patients stratified by treatment. In Patient 1, CRP levels were lower while on treatment with dasatinib monotherapy or dasatinib and etanercept combination therapy. In Patient 2, CRP levels were lower on treatment with etanercept and colchicine combination therapy. Patient 3 had an improvement of the CRP levels and thrombocytopenia on etanercept monotherapy for 3 months; the liver function tests (LFTs), gamma-glutamyltransferase (GGT) and alanine aminotransferase (ALT) did not normalize. **b**. Three liver biopsies obtained from Patient 1 showed bridging fibrosis (upper left and center panels), which improved significantly at 30 months on dasatinib monotherapy (upper right panel). Bile ductopenia recovered (lower left panel) and GGT and ALT levels improved on dasatinib monotherapy or with dasatinib and etanercept combination therapy (lower center and right panels). **c**. Heatmap depicts changes in serum cytokine and chemokine concentrations in longitudinally collected samples from Patient 1 compared to 3 healthy controls. (*) indicates sample obtained when dasatinib was temporarily stopped. **d**. shows heatmap of transcriptome analysis of 4 liver biopsies obtained from Patient 1. A recovery of epithelial markers (e.g. cholangiocytes) and improvement of fibrosis and inflammation markers was observed. Markers of liver sinusoidal endothelial cells (LSEC) activation, of scar/fibrosis associated mesenchymal cells (SAMec) and scar associated macrophages (SAMacs) decreased on dasatinib treatment. ECM, extracellular matrix; HSC, hepatic stellate cells; GC, glucocorticosteroids; IVIG, intravenous immunoglobulin.

Liver sinusoidal endothelial cells (LSEC) express Lyn kinase and are specialized ECs that play a central role in liver homeostasis by regulating intrahepatic vascular tone, immune cell function, and quiescence of hepatic stellate cells (HSCs) ^6-9^. During sustained hepatic injury, LSECs regulate the development of fibrosis^10^ through activation of HSC that differentiate into scar-producing myofibroblasts^11^. Fibrogenic LSEC and HSC signatures have been described in human and murine liver biopsies^11,12^. We analyzed transcriptional signatures associated with liver fibrosis in bulk RNA seq data from four liver biopsies from Patient 1. Markers of fibrogenic LSECs, *CD34*, and *ACKR1* and the NOTCH3-JAG1 axis that regulates HSCs activation were upregulated in biopsies with bridging fibrosis and decreased on dasatinib treatment. Expression of genes of inflammatory cytokines, chemokines, adhesion molecules (CAMs and integrins), extracellular matrix genes and HSC differentiation into mesenchymal cells also decreased. In contrast, gene transcription of epithelial cell markers increased, thus corroborating remodeling during fibrinolysis and recovery from ductopenia (Fig 4d). The effect of active and overexpressed Lyn kinase on HSC activation has been evaluated in a murine model of carbon tetrachloride (CCl4) induced liver fibrosis, that was reverted with Src kinase inhibitor treatment.^13^

The clinical phenotype and the upregulation of β2-integrin function on neutrophils, and of ICAM-1 on skin endothelial cells and in the liver biopsies combined with the treatment response to dasatinib suggested a pivotal role of Lyn GOF mutations in endothelial dysregulation and as driver of vasculitis and liver fibrosis. We generated induced pluripotent stem (iPS) cell-derived endothelial cells (iECs), and genetically corrected (isogenic) iECs from Patient 1, and iECs from 3 healthy controls, and assessed the role of the Lyn kinase mutations on endothelial immune activation, adhesion and endothelial transmigration, thus avoiding Lyn overexpression, which can activate downstream pathways^13^ (Fig. 5a,b). Cytokine (IL-1β) stimulation of mutant iECs resulted in increased and prolonged mRNA expression of ICAM-1 and E-selectin but not of VE-cadherin compared to controls (Fig. 5a); this is corrected in the isogenic iEC control but not with dasatinib treatment (Supplementary Fig. 8). Co-culture of patient but not control neutrophils with mutant or isogenic iECs led to elevated IL-6 levels which was blocked by dasatinib and less by the TNF inhibitor (Fig. 5b, c). IL-8 and CXCL1 were also elevated in these co-cultures. Furthermore, co-culture of healthy control neutrophils with mutant but not isogenic or healthy control iECs led to clustered expression of ICAM-1 beneath areas of firmly adherent neutrophils (Fig. 5d). Neutrophil adhesion and ICAM-1 clusters were significantly blocked with dasatinib and less with the TNF inhibitor at 48 hours (Fig. 5d). To evaluate neutrophil diapedesis, we assessed healthy control neutrophil trans-endothelial migration (TEM); TEM through patient iECs was significantly increased compared to isogenic or healthy control iECs (Fig. 5e) and improved with treatment with dasatinib and the TNF inhibitor (Fig. 5f). In areas of neutrophil adherence to mutant but not healthy control iEC monolayers, VE-cadherin expression was focally reduced, which suggested disruptions of cell–cell contacts and was reflected by the decreased transendothelial electrical resistance (TEER), a measure of endothelial barrier function^14^. TEER significantly improved by TNF inhibition but not dasatinib (Supplementary Fig. 9). These data suggest complementary effects of dasatinib on neutrophil recruitment and adhesion to ECs and TEM, and of TNF inhibition in improving endothelial barrier function, suggesting that both drugs may be effective in improving neutrophilic small vessel vasculitis.^15^

**Figure 5.**
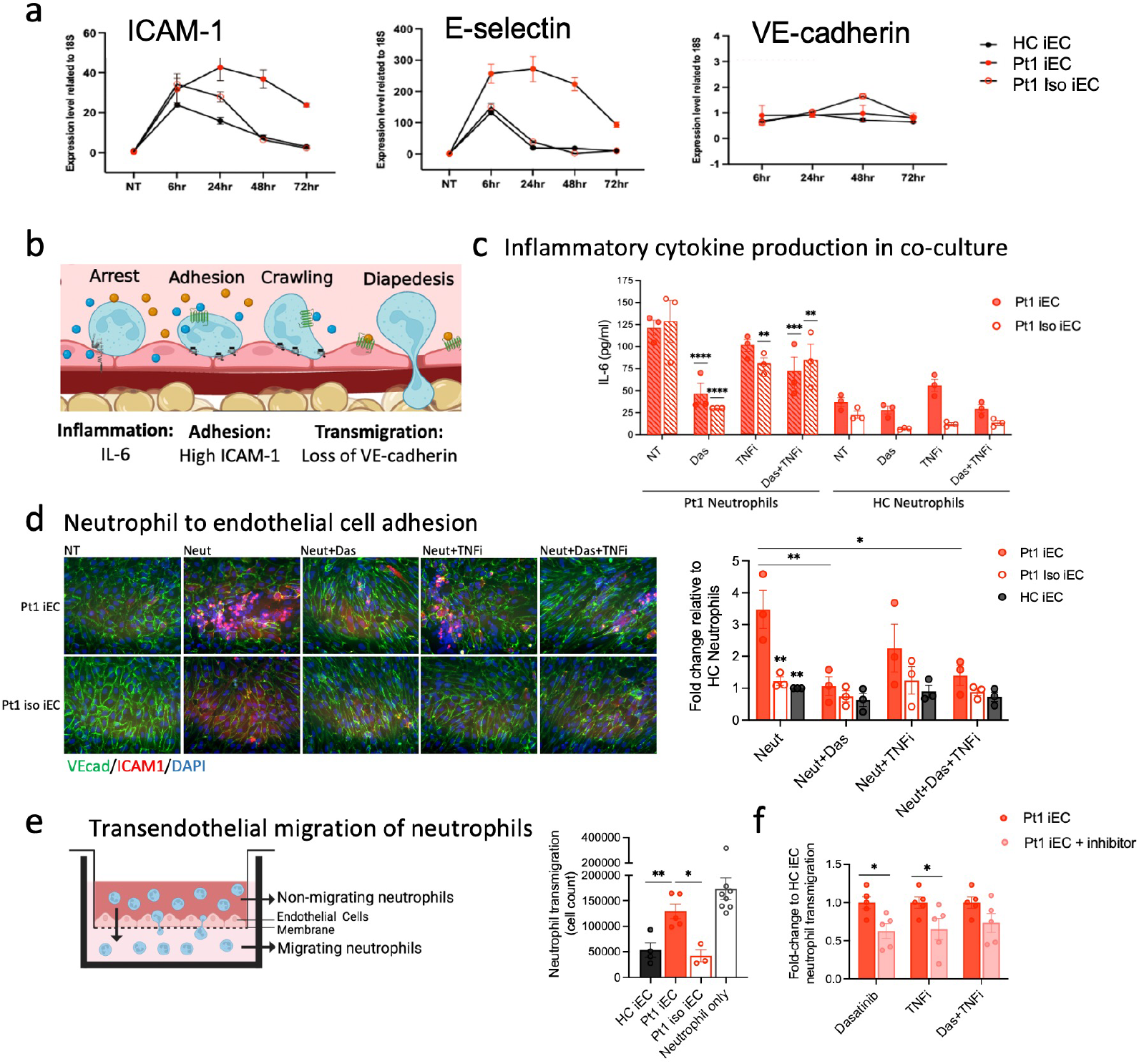
Co-culture of neutrophils and induced endothelial cells (iECs) increases production of inflammatory markers, neutrophil adhesion, and transendothelial migration (TEM). **a**. mRNA expression of the endothelial cell markers ICAM-1, E-selectin, and VE-cadherin (VEcad) in Patient 1’s iECs (Pt1 iECs), a genetically corrected in isogenic iEC clone (Pt1 iso iEC) and a healthy control iEC (HC iEC) was quantified upon stimulation with IL-1β. **b**. Schematic representation of neutrophil extravasation shows critical steps preceding diapedesis. **c**. Under non-flow conditions Pt1 iECs carrying the *LYN* GOF mutation or Pt1 iso iEC were co-cultured with neutrophils from Pt1 or from healthy control for 48 hours. IL-6 production in co-culture supernatant upon treatment with dasatinib (Das),TNF inhibitor adalimumab (TNFi), or both (Das+TNFi) is shown. **d**. Co-cultures of healthy control neutrophils (Neut) with Pt1 iECs (upper panels) or iso IECs (lower panels) for 48 hours lead to significantly increased neutrophil adhesion in areas of dense ICAM-1 expression in mutant compared to iso iEC co-cultures. Treatment with dasatinib (Neut+Das) reduced ICAM-1 expression to control levels. Neutrophil adhesion was quantified after 48 hours. y-axis indicates fold-change of the migrated neutrophil counts to HC iEC and HC neutrophil co-culture counts. Mean of 3 technical replicates from 3 separate experiments are depicted. * p < 0.05, ** p < 0.01, as determined by Two-Way Anova test. **e**. In an endothelial cell transwell migration assay, neutrophil transendothelial migration (TEM) after 24 hours is quantified. * p < 0.05, ** p < 0.01, as determined by Mann-Whitney test. **f**. The effect of Das, TNFi, or both Das + TNFi on TEM of neutrophil migration/diapedesis is depicted; y-axis represents fold-change of the migrated neutrophil counts compared to Pt1 iEC and HC neutrophil co-culture counts. * p < 0.05, as determined by paired t test.

In vitro, the treatment with the pan-Src kinase inhibitor dasatinib does not result in full correction of endothelial and neutrophil dysfunction caused by mutant Lyn, which is however achieved with isogenic gene corrected iECs (Supplementary Fig. 9)^16^. These discrepancies likely reflect the fact that dasatinib indiscriminately blocks the effect of other regulatory Src kinases that are known to collaborate and substitute in regulating endothelial homeostasis^3,17^. Recently, monogenic defects that reduce or eliminate phosphorylation of the conserved inhibitory tyrosine in the C-terminal tail of Src, Hck and Lyn illustrate diverging effects that suggest tissue and organ specificity of Src kinases^5,18,19^. A familial heterozygous GOF *SRC* mutation causes thrombocytopenia, myelofibrosis, bleeding, and non-inflammatory bone pathologies that start in adolescence^20^. In contrast, GOF mutations in *HCK* and *LYN* present with perinatal-onset systemic inflammation, neutrophilic small vessel (or leukocytoclastic) vasculitis, and lung inflammation with a *HCK* GOF mutation and liver fibrosis with *LYN* GOF, respectively. It is therefore intriguing to speculate that engineering of Src-kinase specific inhibitors (i.e. Lyn kinase only) may increase tissue specificity and reduced unwanted effects. Whether Lyn kinase specific inhibition would be a treatment strategy in a wider spectrum of non-syndromic forms of neutrophilic vasculitis will require further evaluations.

In summary, we describe a novel autoinflammatory syndrome, which is caused by *de novo* GOF mutations in *LYN*, the gene that encodes Lyn kinase. We propose to call this syndrome Lyn kinase associated vasculopathy and liver fibrosis (LAVLI). Disease-causing mutations eliminate an inhibitory tyrosine (Y) at position p.Y508 that results in constitutively active Lyn kinase^2^ and a severe clinical phenotype of cutaneous small vessel vasculitis and liver fibrosis. Functional data illustrate a pivotal role of active Lyn kinase in regulating endothelial activation, neutrophil adhesion and transendothelial migration, partly through upregulation of β2 integrins on neutrophils^21,22^, and adhesion molecule expression (i.e. ICAM-1) on endothelial cells^23^. Our study identifies Lyn kinase as potential treatment target^24^ in small vessel vasculitis and early liver fibrosis and illustrates the importance of a genetic diagnosis in patients with recalcitrant inflammatory disease.

## Data Availability

All data produced in the present study are available upon reasonable request to the authors.

## Acknowledgements

The authors would like to thank Rachel Vantries, B.A., Gina Montealegre, M.D., Dawn Chapelle, C.R.N.P., Yan Huang, M.D., April Brundidge, R.N., Alexandra Liptakova, M.D., Anna Sediva, M.D., Ph.D., Milan Macek, Jr, M.D. Ph.D., and Zdenek Sumnik, M.D., Ph.D. for logistical help with patients and sample management; Ondrej Hrusak, M.D., Ph.D., Tomas Kalina, M.D. Ph.D., Milan Macek Jr., MD., Ph.D., Zdenek Sumnik, MD., Ph.D., Anna Sediva, M.D., Ph.D., Kveta Blahova, M.D. Ph.D., Alexandra Liptakova, M.D., Veronika Koukolska, M.D., Quan Yu, M.D, M.S., for their contribution to this study, and the families for their cooperation. Funding was provided by the Intramural Research Program of the NIH, NIAID, NHLBI, NIAMS and the Clinical Center, the Ministry of Health of the Czech Republic (NV19-05-00332, NU20-05-00320, NV18-05-00162 and project for the conceptual development of research organization - 00064203), Ministry of Education, Youth and Sports of the Czech Republic (LM2018132 - to MB and PP) and Charles University (PRIMUS/19/MED/04).

## METHODS

### Study participants

All participants or their legal representatives consented to have the results of this research work published. All research investigations were done as part of protocol 17-I-0016/NCT02974595 and as such were approved by the NIAID and NIAMS/NIDDK Institutional Review Boards. For patient 3, the research investigations were also approved by the Institutional Review Board of the Second Faculty of Medicine, Charles University. Written informed consent was obtained from the parents of all patients/healthy controls involved, and assent was obtained from the patients/healthy controls were indicated.

### Genetic and functional analyses

We performed genetic analyses including whole exome and Sanger sequencing of the patients and their parents. Genomic studies are detailed in the Supplementary Appendix.

## FUNCTIONAL STUDIES

### Phosphorylation Assay of Lyn Kinase and its Substrates

#### Cell Culture

PLB-985 cells were maintained in RPMI 1640 supplemented with 10% fetal bovine serum, 100 units/ml penicillin, and 100 μg/ml streptomycin at a concentration of 0.1–1 × 10^6^ cells/ml. To differentiate the cells, 1.3% DMSO was added to 2 × 10^5^ cells/ml for 6 days. HEK293FT cells (Thermo Fisher Scientific, Waltham MA, USA) were cultured in DMEM supplemented with 10% fetal bovine serum, and antibiotics at 37°C in 5% CO_2_.

#### Plasmid constructs

The plasmid construct encoding wildtype *LYN* in pcDNA3 vector was kindly provided by S. Watson, University of Birmingham, Birmingham, United Kingdom. Plasmids expressing mutant *LYN* (*LYN*, c.1519C>T; *LYN*, c.1523A>T and *LYN*, c.1524C>G) were generated by site-directed mutagenesis (QuikChange™IIXL Site-Directed Mutagenesis kit, Agilent Technologies, Santa Clara, CA, USA) using the following primer pairs: 5’
s-ctgctgctggtattacccttccgtggctg-3′ and 5′-cagccacggaagggtaataccagcagcag-3′ for c.1519C>T; 5′-ctgctgctggaattgcccttccgtggctgt-3′ and 5′-acagccacggaagggcaattccagcagcag-3′ for c.1523A>T; 5′-gctgctgctgctattgcccttccgtggct-3′ and 5′agccacggaagggcaatagcagcagcagc-3′ for c.1524C>G). Successful introduction of the mutations was confirmed by Sanger sequencing. Plasmid encoding SKAP2 with N-terminal FLAG tag was generated by PCR from HL-60 cell line cDNA with these primers: 5’-ctgaattctcccaaccccagcagcacctc-3’ and 5’-atggatcctcaaatatcatacatctccattatgtagg-3’, followed by cloning into EcoRI and BamHI sites of pFLAG-CMV-2 vector and Sanger sequencing.

#### Transfection

Plasmids encoding wildtype or mutant *LYN* or their combination with a plasmid encoding the known Lyn kinase substrate *SCIMP*^25^ and *SKAP2* were co-transfected to the HEK293FT cell line using lipofectamine 2000 transfection reagent (Thermo Fisher Scientific, Waltham MA, USA) according to manufacturer’s instructions.

#### Cell lysis and immunoblotting

24 hours after transfection, HEK293FT cells were lysed in SDS-PAGE sample buffer followed by 15s sonication (amplitude 50, 4710 Ultrasonic Homogenizer, Cole Parmer) and immunoblotting with antibodies to phosphotyrosine (4G10 culture supernatant produced in house), phospho-Src Family Tyr416 (#2101, Cell Signaling Technology, Danvers, MA, USA), phospho-Lyn Tyr508 (#2731, Cell Signaling Technology, Danvers, MA, USA), rabbit polyclonal to Lyn (sc-015, Santa Cruz Biotechnology, Dallas, TX, USA), or monoclonal antibody to SCIMP (NVL-07, produced in house).^25^

#### Viral Infections

Retroviral transduction of all pMX-GFP constructs was performed as follows: Platinum-GP cells were grown to 70% confluency in a 10-cm tissue culture dish and transfected using 10 μg of retroviral DNA and 3 μg VSV-G. The 72-h viral supernatant was collected and added to 1 × 10^6^ PLB-985 cells for 3 days in the presence of 15 μg/ml polybrene. Cells were sorted for equal GFP expression levels using the cell sorter (FACSAria; BD Biosciences, Franklin Lakes, NJ, USA) at the University of Wisconsin flow cytometry facility.

#### Immunoprecipitation and in Vitro Kinase Assay

5×10^6^ differentiated PLB cells were plated per well of 12 well plate coated with 10μg/ml fibrinogen in mHBSS (HBSS, 20 mM HEPES, 0.1% HSA) and allowed to settle for 30min at 37°C. Cells were lysed by adding equal volume RIPA lysis buffer (50 mM Tris-HCl, pH 7.5, 150 mM NaCl, 1% Triton X-100, 2 mM EGTA, 1 mM dithiothreitol, 0.1% SDS, 0.5% sodium deoxycholate, 1mM Na3VO4, 0.2 mM PMSF, 10 μg/ml aprotinin, 5 μg/ml leupeptin). Lysates were cleared by centrifugation at 21K rcf for 10min at 4°C. Protein concentrations were determined by BCA assay for loading of equivalent total protein amounts. GFP proteins were immunoprecipitated with anti-GFP serum (Invitrogen, Waltham, MA, USA) and GammaBind G-Sepharose beads (GE Healthcare, Chicago, IL, USA). The immune complex was washed twice with RIPA buffer and twice with kinase buffer (10 mM Tris-HCl, pH7.5, 5 mM MgCl2). The immunoprecipitate was put in kinase buffer (40 μL) containing 100 μM ATP and 0.5 μg Sam68 (Santa Cruz Biotechnology) at room temperature for 45 seconds with continuous mixing. The reaction was stopped by addition of 5x sample buffer and the immunocomplexes were separated by SDS-PAGE and probed with anti-phosphotyrosine antibody (4G10 Platinum, Millipore, Burlington, MA, USA). To detect SKAP2 phosphorylation, HEK293FT cells transfected in 12-well plates with constructs encoding SKAP2 and LYN mutants as described above were lysed in 250 μl lysis buffer containing 1% n-Dodecyl-β-D-maltoside, 30 mM Tris-HCl pH 7.4, 120 mM NaCl, 2 mM KCl, 2 mM EDTA, 10% glycerol, cOmplete™ EDTA-free Protease Inhibitor Cocktail (Roche), and PhosSTOP™ phosphatase inhibitor cocktail (Roche). Lysates were cleared by centrifugation at 21000 x g for 10min at 4oC. FLAG-tagged SKAP2 was immunoprecipitated with 5 µl anti-FLAG agarose beads (Merck, Sigma-Aldrich) for 2 h at 4°C, followed by elution with 33μl SDS-PAGE sample buffer. The immunoprecipitates were subjected to immunoblotting with anti-phosphotyrosine (4G10) and anti-FLAG M2 antibodies (Merck, Sigma-Aldrich), and the lysates to immunoblotting with antibodies against LYN (Santa Cruz Biotechnology) and GAPDH (Merck, Sigma-Aldrich).

### Serum Cytokine Analysis

Serum was collected from patients 1 and 3 and stored at -80°C.

#### Patient 1

IL-6 and cytokines and chemokines depicted in Figure 4C were measured in 10 serum samples from patient 1 and 3 healthy controls using the Bio-Plex ProTM Human Cytokine 27-Plex and 21-Plex Immunoassays (Bio-Rad, Hercules, CA, USA). Human cytokine standard group I and II were used for the standard curves. The sera were diluted 1:4 with the sample buffer and 50 μl of the diluted serum were used for the assays. All sera were analyzed simultaneously to avoid batch effects. Values below the limit of detection were reset at the limit of detection. Soluble ICAM-1, L-selectin, E-selectin, lipocalin/NGAL, MMP-9, lactoferrin, MPO, and RANTES were measured on customized, magnetic bead-based, multiplex assay (R&D Systems, Minneapolis, MN, USA) according to the manufacturers specifications for standards and dilutions. The magnetic beads were analyzed on Bio-Plex 3D instrumentation (Bio-Rad). Standard curves were analyzed using nonlinear curve fitting and unknowns were calculated based on the derived equation. Samples that exceeded the highest standards were reanalyzed more dilute until the values fell within the range of the known standards. Two control plasma samples and a control sample spiked with a known quantity of each analyte were analyzed on each plate to assess the inter-plate variation and to determine the effect of the biological matrix on the measurement of each analyte. For most analytes, the control samples had <20% variation from plate to plate, and the recoveries were generally >70%.

#### Patient 3

Serum levels of E-selectin, L-selectin, ICAM-1 and lipocalin were detected by ELISA (Abcam, Waltham, MA) in the samples of Patient 3 and 21 healthy controls. The absorbance was measured by ELx800UV absorbance microplate reader (Agilent Technologies). Serum IL-6 level was determined by routine in-house method (chemiluminescence), and the data were acquired using Immulite 2000 XPi (Siemens Healthcare, Erlangen, Germany).

#### Normalization procedure

In order to plot patients 1, 2 and 3’s serum markers IL-6 (Patients 1, 2 and 3), ICAM-1, L-selectin, E-selectin and lipocalin (Patients 1 and 3) on the same graphs, we normalized data using the 2.5^th^ and the 97.5^th^ percentiles for the healthy controls (HC) for the respective assays that were used. Normalized values depicted in the graphs correspond to the analyte concentration minus 2.5%ile of HC, divided by the 97.5%ile minus 2.5%ile of HC, as

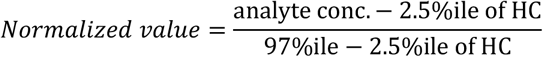

### Neutrophil Characterization and Functional Assessment

#### PMN Surface Antigen Expression

Whole blood samples from healthy controls and patients were collected into EDTA tubes (Vacutainer; BD Biosciences). Aliquots (100 μl) of whole blood were incubated for 15 min at ambient (room) temperature with the following antibodies: CD11b, CD11b (activation epitope) (eBioscience), CD11a, CD11c, CD16, CD18, CD32 CD45, or CD63, CD62L, CD63, CD64 (BD Biosciences). The tubes were treated with OptiLyse C (Beckman Coulter Inc) for 10 min at room temperature to lyse the erythrocytes, washed twice, and then analyzed on the BD FACSCanto II cytometer (BD Biosciences). Expression, as measured by mean fluorescence intensity (MFI), was measured for each surface antigen.

#### PMN Chemotaxis

Neutrophil chemotaxis was measured using an EZ-TAXIScan (Effector Cell Institute, Tokyo, Japan). Isolated neutrophils (1.0 µl of 5×10^6^/ml) were added to the “Cell” well of the EZ-TAXIScan and 1.0 µl of either buffer or *N*-formyl-methionyl-leucyl-phenylalanine (fMLF, 5×10^−8^ M) was added to the opposing “Chemoattractant” well. Digital images of the migrating PMNs were captured every 2.5 min for 1 hr. Images were converted to stacks using the ImageJ software (version 1.52p; NIH). Ten randomly selected cells were electronically traced using the ImageJ plug-in, MTrackJ. The paths of the migrating cells were plotted with the position of each cell at t = 0 anchored at the origin. Using the coordinates of the individual cells in each image, the distance that each cell migrated, and their average velocity, were calculated using the distance formula.

#### PMN Cytokine Stimulation

PMN were isolated from heparinized blood using standard Ficoll-Paque™ Premium (GE Healthcare, Uppsala) discontinuous gradient centrifugation followed by a Dextran (Pharmacosmos, Holbaek, Denmark) sedimentation. The PMN were washed twice with HBSS without divalent cations and resuspended at 2×10^6^/ml in RPMI containing 10% heat-inactivated fetal bovine serum (Thomas Scientific, Swedesboro, NJ). One hundred microliters of the cell suspension were cultured in each well of a Costar #3596 96-well tissue flat-bottom culture plate (Corning, Corning, NY). An equal volume of the following agonists (final concentration) was then added to each well: media alone, lipopolysaccharide (LPS, 200 ng/ml, InvivoGen, San Diego, CA), Phorbol myristate acetate (PMA, 0.2 ng/ml, Sigma Chemical, St. Louis, MO) + Ionomycin (0.5 μM, Sigma Chemical, St. Louis, MO), SAC (0.01%, EMD Chemicals, San Diego, CA), muramyl dipeptide (MDP, 10 μg/ml, InvivoGen), Pam3CSK4 (1 μg/ml, InvivoGen), heat-killed *Listeria monocytogenes* (HKLM,10^8^ cells/ml, InvivoGen), Flagellin (1 μg/ml, InvivoGen), FSL1 (0.1 μg/ml, InvivoGen), ssRNA40 (10 μg/ml, InvivoGen), ODN2006 (5 μM, InvivoGen), thapsigargin (100 nM, Sigma Chemical, St. Louis, MO), opsonized zymosan (1 mg/ml, Sigma Chemical, St. Louis, MO), heat-killed *Candida albicans* (HKCA, 10^8^ cells/ml, InvivoGen, San Diego, CA), depleted zymosan (1 mg/ml, InvivoGen, San Diego, CA), Heparin (40 U/ml, APP Pharmaceuticals, Schaumburg, IL), fMLF, (5×10^−9^ M, Sigma Chemical, St. Louis, MO) + Heparin (40 U/ml), Fibrinogen (2 mg/ml, Sigma Chemical, St. Louis, MO) + Heparin (40 U/ml), fMLF (5 × 10^−9^ M) + Fibrinogen (2 mg/ml) + Heparin (40 U/ml), R848 (5 µM, InvivoGen, San Diego, CA) and ADP Heptose (5 μg/ml, InvivoGen, San Diego, CA). Plates were incubated in a 37°C CO_2_ incubator for 24 hr. After incubation, 4 μl of 10% Triton (Sigma Chemical, St. Louis, MO) was added to wells and the solutions in the wells were mixed using a pipette to disrupt cell membranes. Samples were analyzed using Bio-Plex Pro Human Cytokine 27-plex Assay and Bio-Plex Pro Human IL-8 (BioRad, Hercules, CA).

#### PMN Luminol-Enhanced Chemiluminescence

Isolated PMN (5×10^5^ cells/ml in HBSS with divalent cations and 10 mM HEPES) were transferred to each well a 96-well, white analysis microplate (655207, Greiner Bio-One, Monroe, NC). Luminol ([200 μM]_final_ in HBSS/HEPES) was added to each well of the microplate and the plate was incubated for 10 min at 37°C in the dark. Agonists (buffer, PMA [100 ng/ml]_final_, opsonized zymosan [100 μg/ml]_final_, fMLF [10^−7^ M]_final_, or opsonized heat-killed staph [target:effector ratios of 100:1, 500:1, 1000:1] were added to the wells. The plate was immediately placed in a luminometer (Glomax Multi+, ProMega, Madison, WI) thermally regulated at 37°C. Luminescence was recorded every two min for up to 120 min, shaking the plate at medium speed prior to each reading.

#### PMN Phagocytosis

PMN phagocytosis was assessed using the pHrodo™ *Staphylococcus aureus*

BioParticles™ conjugate for phagocytosis according to the manufacturer’s instructions.

#### Adherence Mediated PMN Superoxide Production

To promote neutrophil adherence, the wells of a 96-well plate (Primaria; BD Biosciences) were coated with fibrinogen by incubating 1 hr at 37°C with 32 μl of 2.5 mg/ml of fibrinogen (EMD Millipore). The wells were then washed 3 times with phosphate-buffered saline. Neutrophils (2×10^5^/200 μl Hanks balanced salt solution with divalent cations/10 mM HEPES/0.2% fetal bovine serum) and cytochrome C (100 μM) were added to the wells in duplicate and preincubated at 37°C for 5 min. An additional identical well that contained superoxide dismutase (100 μg/ml) served as a blank to validate superoxide production. The cells were then treated with buffer alone, tumor necrosis factor α (TNF-α, 50 ng/ml), or phorbol 12-myristate 13-acetate (100 ng/ml). The optical density at 549.5 nm (OD_549.5 nm_) of each well was determined at 5-min intervals for 60 min using a thermally regulated Spectromax Plus 384 plate reader (Molecular Devices) with shaking. The superoxide dismutase blank was subtracted from the duplicate experimental wells, and the relative change in OD_549.5 nm_ vs time was plotted.

#### PMN Adherence

PMN (resuspended at 10×10^6^ cells/ml of HBSS/HEPES (10 mM) with 2% bovine serum albumin (Sigma-Aldrich, St. Louis, MO)) were incubated in the dark with calcein-AM ([5 μg/ml]_final_, Molecular Probes, Eugene, OR) for 15 min at 37°C. The calcein-loaded cells were washed twice with HBSS without divalent cations by centrifugation at 300 x *g* for 5 min. The cell suspension was adjusted to 2×10^6^/ml of HBSS/HEPES and 160 ul of cell suspension was added in triplicate to wells of a 96-well plate that had been pre-incubated for 60 min at 37°C with 32 μl of either HBSS/HEPES buffer, fetal bovine serum (Hyclone), or fibrinogen [2.5 mg/ml]_final_, Sigma-Aldrich). The cells were pre-incubated for 10 min prior to the addition of either buffer or PMA [100 ng/ml]_final_. After an additional 30 min incubation, the well were washed 3x with HBSS/HEPES/BSA to remove non-adherent cells. Finally, 160 μl of HBSS/HEPES/BSA was added to the wells and the fluorescence was determined using a Gemini EM (Molecular Devices, San Jose, CA) using an λ_Ex_ = 494 nm and λ_Em_ = 517 nm. The fluorescence of each well was compared to the fluorescence of wells containing calcein-loaded PMN that had not been treated or washed. The data are expressed % PMN remaining adhered to the well.

#### PMN Granule Proteins in Plasma

Levels of MMP-9, lactoferrin, MPO, and lipocalin-2/NGAL in plasma were measured on customized, magnetic bead-based, multiplex assay (R&D Systems, Minneapolis, MN) according to the manufacturers’ specifications for standards and dilutions. The magnetic beads were analyzed on Bio-Plex 3D instrumentation (Bio-Rad, Hercules, CA). Standard curves were analyzed using nonlinear curve fitting and unknowns were calculated based on the derived equation. Samples that exceeded the highest standards were diluted and reanalyzed until the values fell within the range of the known standards. Two control plasma samples and a control sample spiked with a known quantity of each analyte were analyzed on each plate to assess the inter-plate variation and to determine the effect of the biological matrix on the measurement of each analyte. For most analytes, the control samples had <25% variation from plate to plate, and the recoveries were generally >70%.

### Detection of NETs in Tissue

NETs were detected in tissue as preciously described.^26^ Briefly, tissues were dehydrated with various ethanol dilutions. After antigen retrieval, samples were blocked and incubated with anti-citrullinated histone H4 (Millipore, 1:100) overnight. After a series of washes with PBS, tissues were incubated with Anti-rabbit Alexa Fluor 555 (Life technologies) secondary antibody. Nuclei were counterstained with Hoechst for 10 minutes. Images were acquired on a Zeiss LSM780 confocal laser-scanner microscope.

### Generation of iECs and Isogenic Controls and Functional Studies

The patient-specific induced pluripotent stem cells (iPSC) were generated from peripheral blood cells of patient via activation of Yamanaka’s transcription factors (Oct4, Sox2, Klf4, and C-Myc). By using CRISPR-Cas9 gene editing technology, we obtained the isogenic control iPSC line originating from patient-specific iPSC. We have also derived iPSC lines from healthy volunteers (control) who did not have the *LYN* mutation. All iPSC lines tested had the potential of unlimited self-renewal and the ability of three germ layers differentiation (data not shown). The iPSC lines were differentiated into endothelial cells (iEC) following our previously published protocol.^27^ In brief, iPSCs were seeded in Matrigel coated plates at low density supplement with TeSR-E8. After 24 hours, culture cells were induced toward mesoderm progenitors in mesoderm differentiation medium for 6 days. Consequently, the CD31 positive cells from the induction culture were enriched by using CD31 magnetic beads, and further cultured and expanded in Collagen I coated plates or dishes with endothelial culture medium. For immunostaining, cells were fixed with 4% paraformaldehyde (PFA) and stained using antibodies for CD31, CD144, and von Willebrand factor (vWF). Nuclei were visualized with DAPI (Thermo Fisher Scientific). Stained cells were photographed with a fluorescence microscopic system (Zeiss, Oberkochen, Germany)

A *low density lipoprotein (LDL) uptake* assay was performed as described previously.^27^ Briefly, the iECs were incubated with 10 μg/ml acetylated LDL (Ac-LDL) labeled with 1,1’-dioctadecyl-3,3,3’,3’-tetramethylindo-carbocyanine perchlorate (DiI-Ac-LDL) (Invitrogen, Catalog# L3484) for 4 h at 37°C. The DiI-Ac-LDL uptake was assessed and quantified by fluorescent microscopy. Subsequently, cells were dissociated into single cells with TrypLE TM Express Enzyme. The data acquisition was performed on a MACSQuant Flow Cytometer (Miltenyi Biotec, Bergisch Gladbach, Germany) and the results were analyzed with FlowJo software (FlowJo, LLC).

#### Co-culture assays

##### Transmigration assay

The transwell inserts of a 24-well plate were coated with fibronectin (31.25 µg/mL) for 30 min and followed seeding iPS-derived endothelial cells (iECs) at density of 60000 cells/well in the upper insert. After iECs formed a confluent monolayer (24–48 h later), freshly isolated healthy control neutrophils at neutrophils/iECs 10:1 ratio were added to the top insert with 1:1 ratio mixed media (iEC media/RPMI with 10% FBS); the same media was added to the bottom chamber. The neutrophils that transmigrated to the bottom chamber were collected at 24 h after co-culture. Cells were counted with Bio-Rad cell counter.

##### Neutrophil adhesion assay

iECs in passage 3 were used for experiments. Cells were co-cultured with or without fresh isolated healthy control or Patient 1 neutrophils at neutrophils/iECs 10:1 ratio for 48 hours then fixed with 4% paraformaldehyde for 10 minutes at RT. The primary antibody VE-cadherin (1:100 Santa Cruz) and ICAM1 (1:100, Abcam) were applied overnight at 4°C, followed by the corresponding secondary antibody for one hour at room temperature. DAPI (1:1000) was used for cell nuclei detection. Cells were imaged using a Zeiss inverted fluorescence microscope. Counting of adhered neutrophil nuclei via DAPI staining for adhesion was conducted on 10 random fields of view of 6 (Pt1 iso iEC & HC iEC) to 9 (Pt1 iEC) technical replicates conducted in 4 separate experiments. Co-culture supernatant was analyzed by ELISA (R&D Systems, Minneapolis, MN) for IL-6 and by proteome profiler human cytokine array (R&D Systems, Minneapolis, MN, USA) for IL-8 and CXCL1.

### Tissue Immunofluorescence Staining

Formalin-fixed, paraffin-embedded skin samples were deparaffinized with two 30 min washes in xylene, then a series of two 100% ethanol washes for 1 minute each, and 1 min washes in 90%, 80%, 70%, 50% ethanol then water. Antigen retrieval was performed in a solution of 10 mM sodium citrate (pH 6.0) or 1 mM EDTA buffer (pH 8.0) for 40 min at 95°C. Once cooled to room temperature (RT) samples were blocked for one hour in 5% BSA, 20% donkey serum and 0.1% Triton-X100 in PBS. Primary antibodies for CD31 (1:50, DAKO, Glostrup, Denmark), E-selectin (1:25, Abcam), ICAM-1 (1:50, Abcam) and vWF (1:200, DAKO) were applied overnight at 4°C, followed by the corresponding secondary antibody for one hour at room temperature. Slides were mounted using DAPI-containing mounting media (Vector Laboratories). Slides were imaged using a Zeiss LSM 510 META confocal microscope.

### Western Blot

Western blot samples were lysed in CHAPS buffer containing protease and phosphatase inhibitors (Invitrogen). For each sample, 20μg of total protein was analyzed by Western blot (Bio-Rad 4–20% Mini-PROTEAN TGX Gel) and transferred onto nitrocellulose membranes. After blocking, membranes were incubated with primary antibody ICAM-1 (1:1000, CST), VE-cadherin (1:1000, CST), CD31 (1:1000 DAKO) and GAPDH (1:1000, CST) overnight at 4°C. Secondary fluorescence antibody (1:10000 LI-COR) was applied for 1h at room temperature. After wash with PBST three times, the signal was detected and quantitated by Odyssey imagining system.

### Electric Impedance Spectroscopy (EIS)

#### Transendothelial Electrical Resistance (TER)

The barrier function of endothelial cells was measured using Electric Cell-substrate Impedance Sensing (ECIS) technology (Applied BioPhysics, Troy, NY, USA). The ECIS arrays (8W10E+) were pretreated with 10 mM L-cysteine, followed by coating with fibronectin (Sigma). The iEC were seeded at 120,000 cells per well in 400 μL of iEC growth medium. The barrier integrity of iEC was monitored for 48 h when the cells were treated with dasatinib (10nM, and 50nM, Selleckchem, Houston, TX), TNFα inhibitor adalimumab (12μg/ml), and dasatinib plus TNFα inhibitor. The multiple frequency data were collected for barrier modelling as recommended by Applied Biophysics. ECIS measurements were acquired from three independent biological repeats with three replicates per experiment. The data shown were from one independent experiment with three replicates, which is representative of the three biological repeats. The results were graphed using GraphPad Prism software (GraphPad Software).

### Phosphorylation Assay of Lyn Kinase and Downstream Substrates in Patient B cells

Freshly isolated PBMCs were treated with 50 mM/ml PP2 (Sigma-Aldrich, St Louis, MI, USA) or 100 nM/ml dasatinib (Selleckchem, Houston, TX) or DMSO for 1h at 37°C, then stimulated with 10 mg/ml goat F(ab’)_2_ anti-human IgM (polyclonal; Jackson ImmunoResearch Laboratories) at 37°C for 2 min. For the detection of phosphorylated signaling intermediates, cells were fixed and permeabilized using BD Cytofix and Phosflow Perm/Wash buffers (BD Biosciences, Franklin Lakes, NJ, USA), according to manufacturer instructions and stained separately with PE-conjugated mouse anti-human antibodies against phosphorylated Syk (p-Y348) and PLC-g2 (p-Y759) (BD Biosciences), or unconjugated rabbit anti-human antibodies against phosphorylated Lyn (p-Y397), CD19 (p-Y530), CD22 (p-Y822) (Abcam, Cambridge, UK), CD79A (p-Y182) (Cell Signaling Technology, Danvers, MA, USA) followed by staining with secondary goat anti-rabbit IgG-PE (Thermo Fisher Scientific, Waltham, MA, USA). PerCP-Cy5.5-conjugated mouse anti-human CD19 (Thermo Fisher Scientific) was included in each stain. The cells were acquired on a FACSCanto II (BD Biosciences) and analyzed using FlowJo software (BD Biosciences).

### Macrophage Stimulation Assay and Cytokine/chemokine production

Adherent monocytes were seeded in multi-well plates or dishes and were differentiated into M1-like and M2-like monocyte-derived macrophages (M1-MDMs and M2-MDMs) by culturing with RPMI 1640 supplemented with 10% FBS containing either 10 ng/ml recombinant (rh) GM-CSF, or 100 ng/ml recombinant human (rh) M-CSF (Peprotech, USA) for M1-, and M2-MDMs respectively. Macrophages were stimulated in the presence of lipopolysaccharide (LPS) (1 μg/ml) and ATP (1 mmol) (Sigma-Aldrich, St. Louis, MO, USA). Culture supernatants for macrophages were collected after 24 hours. Relative levels of multiple cytokines and chemokines in the supernatants of macrophages were analyzed using a proteome profiler human cytokine array (R&D Systems, Minneapolis, MN, USA) according to the manufacturer’s instructions. In brief, culture supernatants (200 μl) of macrophages, which were collected after centrifugation, were added to dot blots onto which the capture antibodies had been spotted in duplicates. After incubation with the secondary antibody mixture, the resultant signals were detected using the Bio-Rad (Hercules, CA, USA) image analyzer. The intensity of the spots was quantified using the ImageJ software.

### STAT Phosphorylation Assay in Patient Monocytes

Cryopreserved PBMCs were assessed for signal transducer and activator of transcription (STAT) phosphorylation. For the detection of phosphor-STAT1 (Y701; BD 612596) and pSTAT6 (Y641;BD 612600), PBMCs (1X 10^6^) were stimulated with or without IFNa (10 ng/ml; Cell signaling 8927), IFNg (10 ng/ml; Peprotech 300-02), or IL-4 (10 ng/ml; Peprotech 200-04) for 20 minutes at 37°C. Cells were fixed using BD Cytofix Fixation Buffer (15 minutes at 37°C) and then permeabilized in BD Phosflow Perm Buffer III (30 minutes on ice) according to manufacturer’s instructions. Cells were then washed and stained with phospho-STAT (BD bioscience) antibody in the stain buffer for one hour. The cells were acquired and analyzed by a BD FACSCanto II flow cytometry and Flow Jo (Treestar), respectively.

### qRT-PCR Quantification of *LYN* Gene Expression

Total RNA was extracted from different types of cells and tissues by using TRIzol (Thermo Fisher Scientific, Waltham, MA), and the quantification of total RNA was performed using the 2100 Bioanalyzer (Santa Clara, CA). Reverse transcription was performed using the High-Capacity cDNA Reverse Transcription Kit (Applied Biosystems, Waltham, MA) to generate the first strand synthesis cDNA. The Lyn kinase encoding gene (*LYN*) messenger RNAs was quantified by qRT-PCR using Taqman Gene Expression Assay (Hs01015816_m1, Applied Biosystems, Waltham, MA, USA) and normalized against 18S gene expression. The Lyn mRNA relative expression obtained from the different cell and tissue types are reported as mean fold-change (+/-SEM) and further normalized against whole blood sample (blue dotted line).

### Analysis of Lyn Protein Expression by Western Blot in Tissues and Cell Subsets

Lyn protein expression was measured using Western blot. Healthy donor whole blood (age range 18-50 years) was obtained from the NIH Blood Bank (Bethesda, MD), and was used to isolate total PBMCs and neutrophils. These cell type populations were isolated using ficoll (Histopaque Ficoll, Sigma-Aldrich/Millipore St. Louis, MO), and separated using density-gradient centrifugation. The PBMCs were stained with fluorescently labeled antibodies that bind to specific epitopes such as CD14+ (Monocytes), CD3+ (T cells), CD19+ (B cells), CD56/CD16+ (NK). Once stained, the viable population of cells were sorted by flow cytometry (FACSCanto II, BD Biosciences) to isolate monocytes, T, B and NK lymphocytes. In addition, biopsied and cell-cultured skin fibroblasts and keratinocytes, and commercially available liver tissue and endothelial cells were. The different cell types were lysed using RIPA buffer (Cell Signaling, Danvers, MA), supplemented with 1 mM PMSF and protease inhibitor cocktail (Roche Mannheim, Germany). From the cell lysate generated, protein concentration was determined using the BCA Protein Assay Kit (Pierce-Thermo Scientific, Pittsburg, PA). For analysis of Lyn protein expression, 15μg of total cell lysate was loaded and separated using a 10% NuPAGE Tris Bis gel under reducing and denaturing conditions, and then transferred to nitrocellulose (Left WB Panel) and PVDF (Right WB Panel) membranes. The membranes were blocked with either 5% milk or odyssey blocking buffer (LI-COR, Lincoln, NE), blotted with purified-unconjugated anti-Lyn Ab (MAB3206 1:500 R&D Systems, Minneapolis, MN), and followed by the IRDye secondary antibody (1:10,000 800CW and 680RD LI-COR). The membranes were scanned with an Odyssey Scanner (LI-COR). Equal protein loading was based on the total protein lysate quantification using the BCA assay and the Beta-Actin expression amongst the different cells and tissues. Positive protein expression of Lyn was identified by double bands detected at 56kDa and 53kDa.

