## Supplementary Appendix for "Constitutively active Lyn kinase causes a cutaneous small vessel vasculitis and liver fibrosis syndrome"

This research was supported by the Intramural Research Program of the NIH, NIAID, NHLBI, NIAMS, and the Clinical Center.

#### CONTENTS

#### I. Supplementary Methods

1. GENOMIC STUDIES
2. FUNCTIONAL STUDIES
3. ASSESSMENT OF CLINICAL RESPONSE

#### II. Supplementary Figures

1. Supplementary Figure 1. WES Filtering for de novo Variants, Sanger Sequencing Validation, and Lyn Protein Multi Species Alignment.
2. Supplementary Figure 2. Presence of Neutrophils in Areas of Small Vessel Vasculitis that Stain Positive for Lyn Kinase.
3. Supplementary Figure 3. Expression of Lyn, Other Src Kinases and Selected Adapter Molecules in Several Cell Subsets and Tissues.
4. Supplementary Figure 4. Phosphorylation Assay of Lyn Kinase and Downstream Substrates in Patient B Cells.
5. Supplementary Figure 5. M1-Like Monocyte-Derived Macrophages (MDM) Stimulation Assay and Monocyte STAT Phosphorylation in Patient 1.
6. Supplementary Figure 6. Effect of Various Treatments on Neutrophil Activation.
7. Supplementary Figure 7. Treatment Effect on Biliary Ductopenia and Liver Fibrosis in Patient 1.
8. Supplementary Figure 8. Lyn Mutations Increase Neutrophil Activation and Neutrophil Transendothelial Migration (TEM).
9. Supplementary Figure 9. Assessment of Vascular Permeability by VE-Cadherin Staining and by Transendothelial Resistance by Electric Cell-Substrate Impedance Sensing (ECIS).

#### III. Supplementary Tables

1. **Supplementary Table 1: Features of the *LYN* Mutations Detected in Patients 1, 2 and 3**
2. **Supplementary Table 2: Clinical and Laboratory Features of Patients with *LYN* Gain-of-Function Mutations**
3. **Supplementary Table 3: Broad Assessment of Neutrophil Function**
4. **Supplementary Table 4: Summary of Liver Biopsies with Progression of Liver Disease Prior to Dasatinib Treatment**

#### IV. Supplementary Results

1. **Supplementary Results: Evaluation of autoimmune dysregulation**

    i. **Supplementary Figure i: Assessment of B Cell Function in Bone Marrow and Peripheral Compartment.**
    ii. **Supplementary Figure ii: Evaluation of Treg Cell Function in Patient 1 Compared to a Healthy Control.**
    iii. **Supplementary Figure iii: Spleen Histology of Patient 1 with Splenectomy**

#### V. Supplementary References

### I. Supplementary Methods

#### 1. GENOMIC STUDIES

**Next Generation Sequencing (NGS):** Whole Exome Sequencing (WES) was performed in patient 1 and his parents' DNA samples. Agilent SureSelect® Human 51 Mbp All Exon Kit (Agilent Technologies, Santa Clara, CA) was used for exome capture. Sequencing was performed on Illumina® HiSeq sequencers (Illumina, San Diego, CA) using 2x100 bp paired reads. The typical average on-target coverage is about 66X. A computational pipeline was developed to process the read data and perform tasks such as quality control (QC), variant discovery, annotation and filtering. Briefly, the sequencing reads in FASTQ format were aligned to the human reference genome (GRC B37) with the BWA (Burrows-Wheeler Aligner) mapping tool. The resulting BAM files (one per sample) were further processed to remove duplicate reads, refine alignment around indels and recalibrate base quality scores, according to the best practice guideline by the Genome Analysis Toolkit (GATK) from the Broad Institute. A variant caller (UnifiedGenotyper) from GATK was used to make joint variant calls across multiple samples, followed by a variant quality score recalibration step by the GATK VQSR tool. Besides quality scores, the variants were annotated with functional impact and allele frequency in public databases and local datasets. Sample relationship and gender were checked to identify potential sample mix-up and false family relationship. Likely disease-causing mutations were selected and prioritized based on quality score, allele frequency, functional impact, probable inheritance model (de novo mutation, autosomal recessive, autosomal dominant, or X-linked), and expert evaluation. A targeted NGS gene panel was used for the identification of the *LYN* mutation in patient 2, and patient 3's mutation was identified by Sophia Clinical Exome Solution (Sophia Genetics, Geneva, Switzerland) using SOPHiA DDM bioinformatics platform (human reference genome GRCh37). The presence of detected variant was validated by Sanger sequencing, and *de novo* occurrence was proved through subsequent family segregation analyses by Sanger sequencing.

**Sanger Sequencing**: The reference sequence used for primer design and nucleotide numbering was *LYN* NM_002350. Exon 13 and its flanking intronic sites were amplified by polymerase chain reaction (PCR) using specific primers (IDT, Coralville, IA) designed with Exon Primer* or Primer-BLAST**. Two options of primers are listed below:

| Exon | Primer sequence | PCR product size (bp) |
|---|---|---|
| Exon 13_option 1 | Forward - TGCCAGGTTTCTAAACGG | 339 |
| | Reverse - CATAAGTGCAACCAGTGATGG | |
| Exon 13_option 2 | Forward – TGTGAGGAGCAAAAAGACCACT | 402 |
| | Reverse – GGCAGATGATCCCCGCACA | |

*Exon Primer: http://ihg.gsf.de/ihg/ExonPrimer
**Primer-BLAST: https://www.ncbi.nlm.nih.gov/tools/primer-blast

**Gene-Expression Analysis by RNA Sequencing (RNA-Seq)**:
Total RNA was extracted from four liver biopsies obtained from patient 1. As a control, a commercially available RNA sample extracted from human liver (OriGene, Rockville, MD, USA) was used. Total RNA integrity was analyzed with Agilent 2100 Bioanalyzer. mRNA purification and fragmentation, complementary DNA synthesis and target amplification were performed using the Illumina® TruSeq RNA Sample Preparation Kit (Illumina, San Diego, CA, USA). Pooled cDNA libraries were sequenced using HiSeq 2000 Illumina® platform (Illumina). Sequencing results were analyzed using Partek GS v6.6 software and are expressed in RPKM

(reads per kilobase exon per million mapped. All RPKM values were offset by the addition of 0.5 prior to analysis to limit the effects of low-expressing samples.[1]

#### 2. ADDITIONAL FUNCTIONAL STUDIES

*Gene Expression by RT-qPCR*

The iEC were seeded and cultured into collagen I coated 12-well plates at the density of 100,000 cells per well for overnight. Followed by treatment with IL-1β, total RNA of iEC was isolated by using RNeasy Mini Kits (Qiagen, Hilden, Germany). cDNA was synthesized by reverse transcription (RT) using Super Script™ III (Invitrogen, Waltham, MA, USA). RT-qPCR was performed using SYBR Green Premix on a Real-Time PCR Detection System (Bio-Rad). Assays were run in duplicate, and results were normalized to 18S ribosomal RNA expression. Primers used for RT-qPCR are shown in the table below.

Primers used for RT-qPCR and PCR

| Target | Forward/reverse primer (5'-3') |
|---|---|
| *ICAM* | TCT GTG TCC CCC TCA AAA GTC/ GGG GTC TCT ATG CCC AAC AA |
| *E-selectin* | ACC TCC ACG GAA GCT ATG ACT/ CAG ACC CAC ACA TTG TTG ACT T |
| *VE-Cadherin* | AGC CCA AAG TGT GTG AGA ACG C/ CTG AGA TGA CCA CGG GTA GGA A |

#### 3. ASSESSMENT OF CLINICAL RESPONSE

*No response*: ongoing frequent flares of skin disease, stable but abnormal or worsening of liver disease with worsening of fibrosis.

*Partial response*: significantly improved rash, fever and CRP, improved but still elevated LFTs, stable elastography.

*Good response*: no fever, frequency of rash below once per month, normalization of LFTs, improved or normalized elastography.

*Long-term remission:* no fever, frequency of rash below once per month, normalization of LFTs, improved or normalized elastography since the last physician/NIH visit.

*Adverse events*, particularly infections, were captured on the natural history protocol.

#### II. Supplementary Figures

##### Supplementary Figure 1. WES Filtering for *de novo* Variants, Sanger Sequencing Validation, and Lyn Protein Multi Species Alignment.

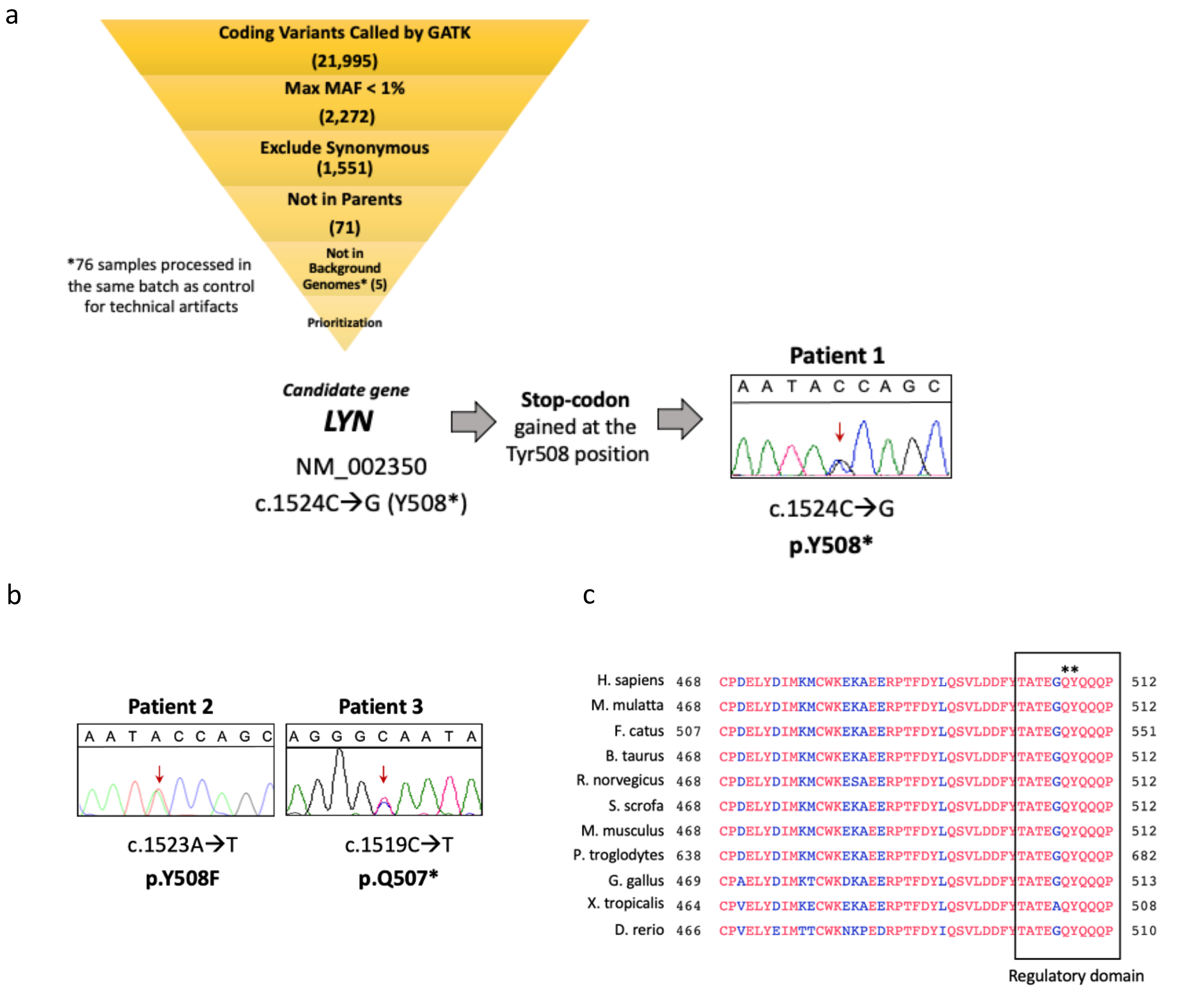


**a.** Filtering process for the detection of *de novo* variants in Patient 1's trio whole exome sequencing (WES) revealed *LYN* de novo variant c.1524C>G, p.Y508* as the best candidate. **b.** Sanger sequencing confirming mutations in Patient 2 and 3, that were identified by a next generation sequencing (NGS) gene panel and WES as having *LYN* c.1524C>G, p.Y508F missense variant and a *LYN* c.1519C>T, p.Q507* truncation respectively. The 3 mutations lead to loss of p.Y508 regulatory tyrosine that, when dephosphorylated, renders Lyn active. The *LYN* mutations were validated by Sanger sequencing in the 3 patients; they were novel, predicted to be deleterious and occurred in highly conserved amino acids. Deleteriousness predictions of the 3 *LYN* variants are detailed in Supplementary Table S1. GATK, genome analysis toolkit; Max, maximum; MAF, minor allele frequency. **c.** Multispecies alignment depicting the position of the 3 Lyn variants (asterisks). The 3 variants affect the protein regulatory domain (black box) and, while Patient 2 has a missense variant (p.Y508F), the variants found in Patients 1 (p.Y508*) and 3 (p.Q507*) result in the loss of p.Y508 and of 4 C-terminal amino acids (QQQP) that are highly conserved among species.

#### Supplementary Figure 2. Presence of Neutrophils in Areas of Small Vessel Vasculitis that Stain Positive for Lyn Kinase.

a

b

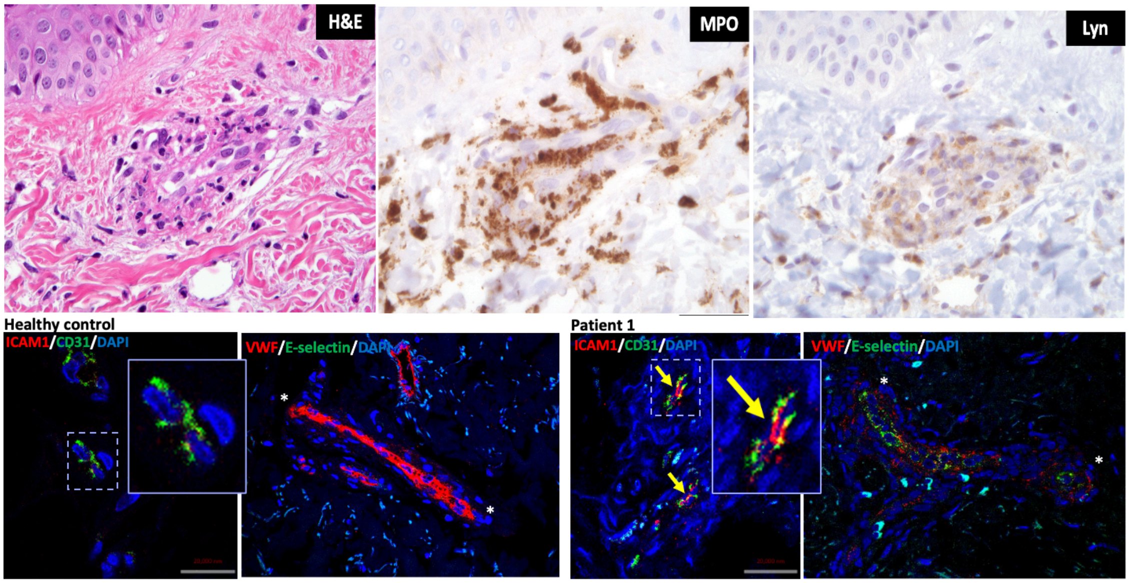


**a.** Histopathology slides depict Patient 1 lesional skin biopsy. Inflammatory infiltrate destroying a vessel wall (H&E, left panel); the inflammatory infiltrate is predominantly composed of neutrophils (MPO+ cells) (center panel) and Lyn staining is present in neutrophils and endothelial cells (right panel). **b.** Compared to healthy controls (left two panels), Patient 1 skin biopsy (right panels) shows loss of vWF staining and focal expression of E-selectin. Asterixis (*) indicate upper and lower pole of vessel.

Not shown: Direct immunofluorescence (DIF) was negative for IgG, IgM and IgA, with weak superficial blood vessels staining of C3. Few superficial blood vessels had patchy staining with fibrinogen. The weak vascular staining was nonspecific and insufficient for a diagnosis of vasculitis. There was no specific evidence for lupus erythematosus, a lichenoid tissue reaction or immune-bullous disease.

#### Supplementary Figure 3. Expression of Lyn, Other Src Kinases and Selected Adapter Molecules in Several Cell Subsets and Tissues.

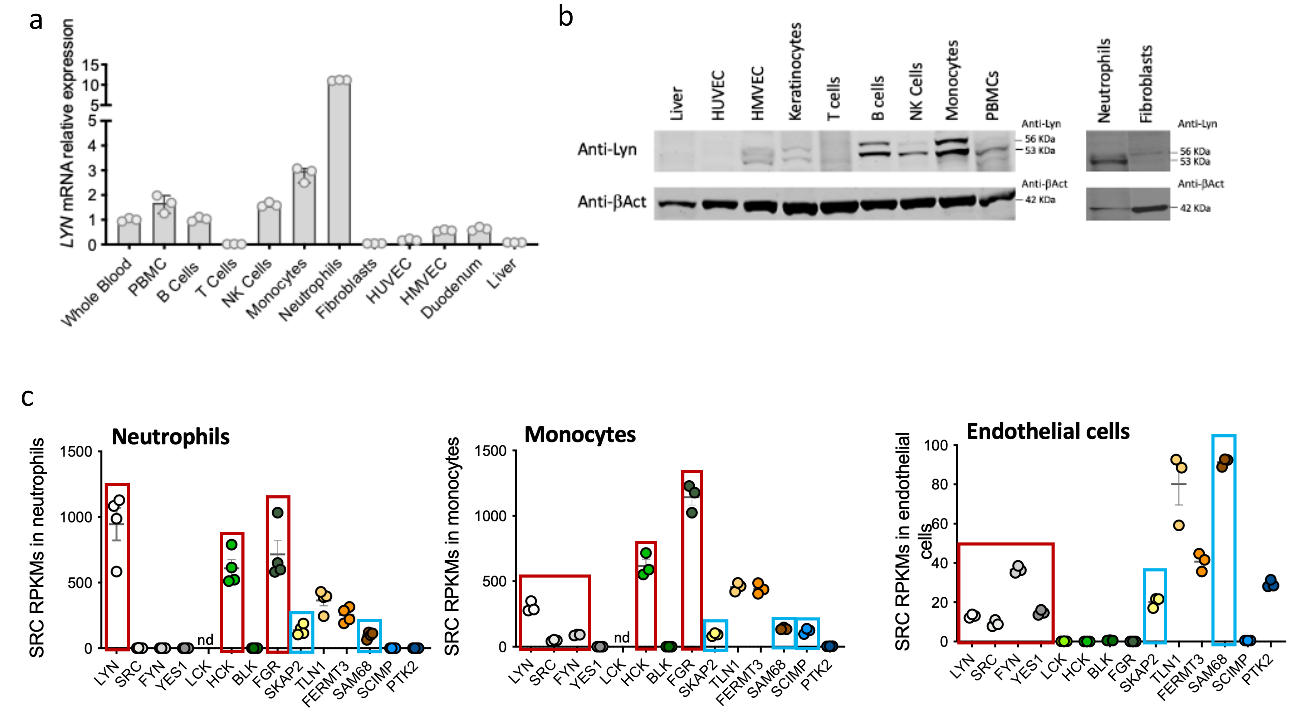


| | Neutrophils | | Refs | Monocytes | | Refs | Endothelial cells | | Refs |
|---|---|---|---|---|---|---|---|---|---|
| **Src** kinase | **Lyn, Fgr, Hck** | Src family members regulate adhesion, activation and migration | Futosi K, Imm Rev 2016 | **Fgr, Hck, Lyn, Fyn, Src** | Regulation of an inflammatory environment | Kovács et al. JEM 2014 | **Fyn, Lyn, Yes1, Src** | Src family members regulate endothelial permeability | Kim MP CTR 2009 |

**a.** Quantitative real-time PCR (RT-qPCR) depicting *LYN* mRNA relative expression to whole blood (WB) in healthy control cell subsets and tissues. **b.** Western blot analysis of Lyn protein expression in healthy control cell subsets and tissues. The anti-Lyn antibody detects the two Lyn isoforms, LynA, 56kDa and LynB, 53kDa. PBMC: peripheral blood mononuclear cells, NK: natural killer, HUVEC: human umbilical vein endothelial cell, HMVEC: human microvascular endothelial cell. **c.** RNA-seq shows that Lyn kinase is most highly expressed in neutrophils, which also express the myeloid-specific Src kinases family members, Hck and Fgr (*red boxes*). Monocytes express lower Lyn kinase levels, low levels of Src and Fyn and similar levels of Hck and Fgr. Lyn expression is lowest in endothelial cells, which also express Src kinases Src, Fyn and Yes1 and lack the myeloid specific kinases. Expression levels of three Lyn-regulated adaptor molecules Skap2, Sam68 and Scimp (*blue boxes*) are also shown; Skap2 and Sam68 are expressed in myeloid and endothelial cells, while Scimp is only expressed in monocytes. RNA-seq was performed on healthy controls' neutrophils (n=4), monocytes (n= 3) and a cultured human lung endothelial cell line (HLMEC) n= technical replicates of 3 different cultures.

The adaptor molecules Scimp, Skap2 and Sam68 are critical in linking active Lyn kinase to downstream functions. Scimp, a TLR adaptor, is only expressed in monocytes and is required for tyrosine phosphorylation of TLR4, and production of proinflammatory cytokines IL-6 and IL-12p40.[2] Skap2 has a crucial role in regulating actin polymerization and binding of talin-1 and kindlin-3 to the β2 integrin cytoplasmic domain and is indispensable for β2 integrin activation and neutrophil recruitment.[3] Its activation is also required for podosome formation and monocyte polarization.[4] SAM68 (Src-associated in mitosis 68 kDa) augments TNFα–induced expression of TNFα, IL-1β, and IL-6 and increases nuclear phospho-p65, and NF-κB activation.[5] Also known as KHDRBS1 (KH domain containing, RNA binding, signal transduction associated 1) it is a member of the STAR family of RNA-binding proteins, which allows the binding of certain RNA sequences with high affinity and regulates protein translation.[6]

#### Supplementary Figure 4. Phosphorylation Assay of Lyn Kinase and Downstream Substrates in Patient B Cells.

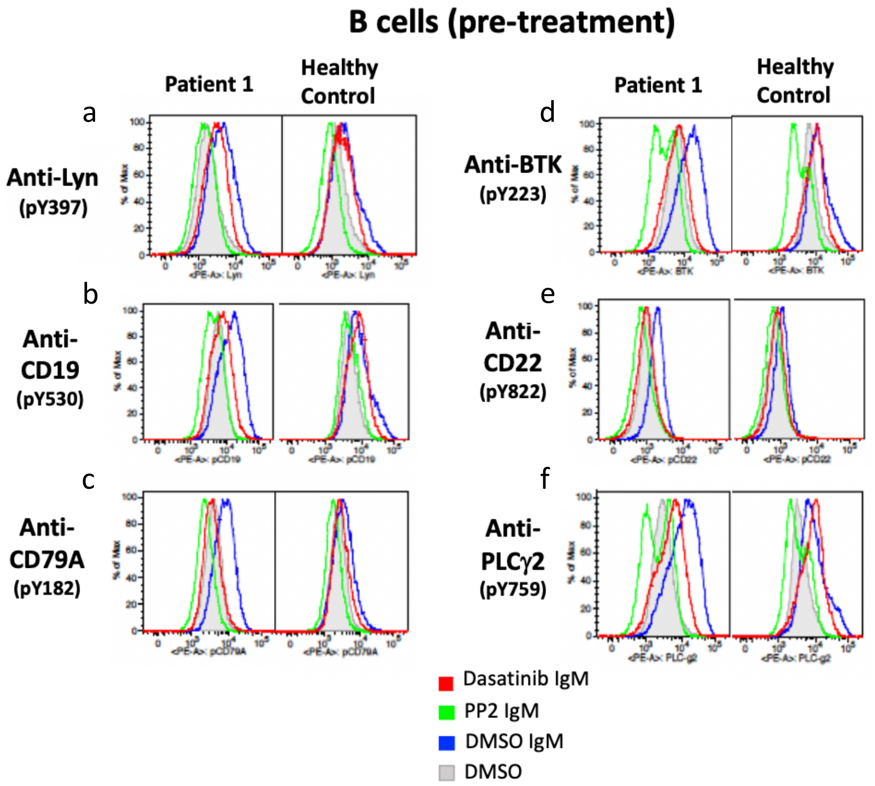


**a-f.** Patient 1 and healthy control PBMCs were stimulated with anti-IgM in the presence and absence of the Src kinase inhibitors PP2 and dasatinib. B cells were assessed by flow cytometry **a.** for autophosphorylation of the $Tyr^{397}$ at the activation loop, **b-f.** and for phosphorylation of Lyn kinase substrates, **b.** phosphorylation of CD19 at position $pTyr^{530}$, **c.** CD79A at position $Tyr^{182}$, **d.** BTK at position $Tyr^{223}$, **e.** CD22 at $Tyr^{822}$, **f.** PLCγ2 at$Tyr^{759}$. IgM B cell receptor (BCR) stimulation of B cells resulted in higher Lyn kinase phosphorylation in patient than control (blue line). Increased phosphorylation was also seen in all Lyn substrates evaluated and was

**Supplementary Figure 5. M1-Like Monocyte-Derived Macrophages (MDM) Stimulation Assay and Monocyte STAT1 and STAT6 Phosphorylation in Patient 1.**

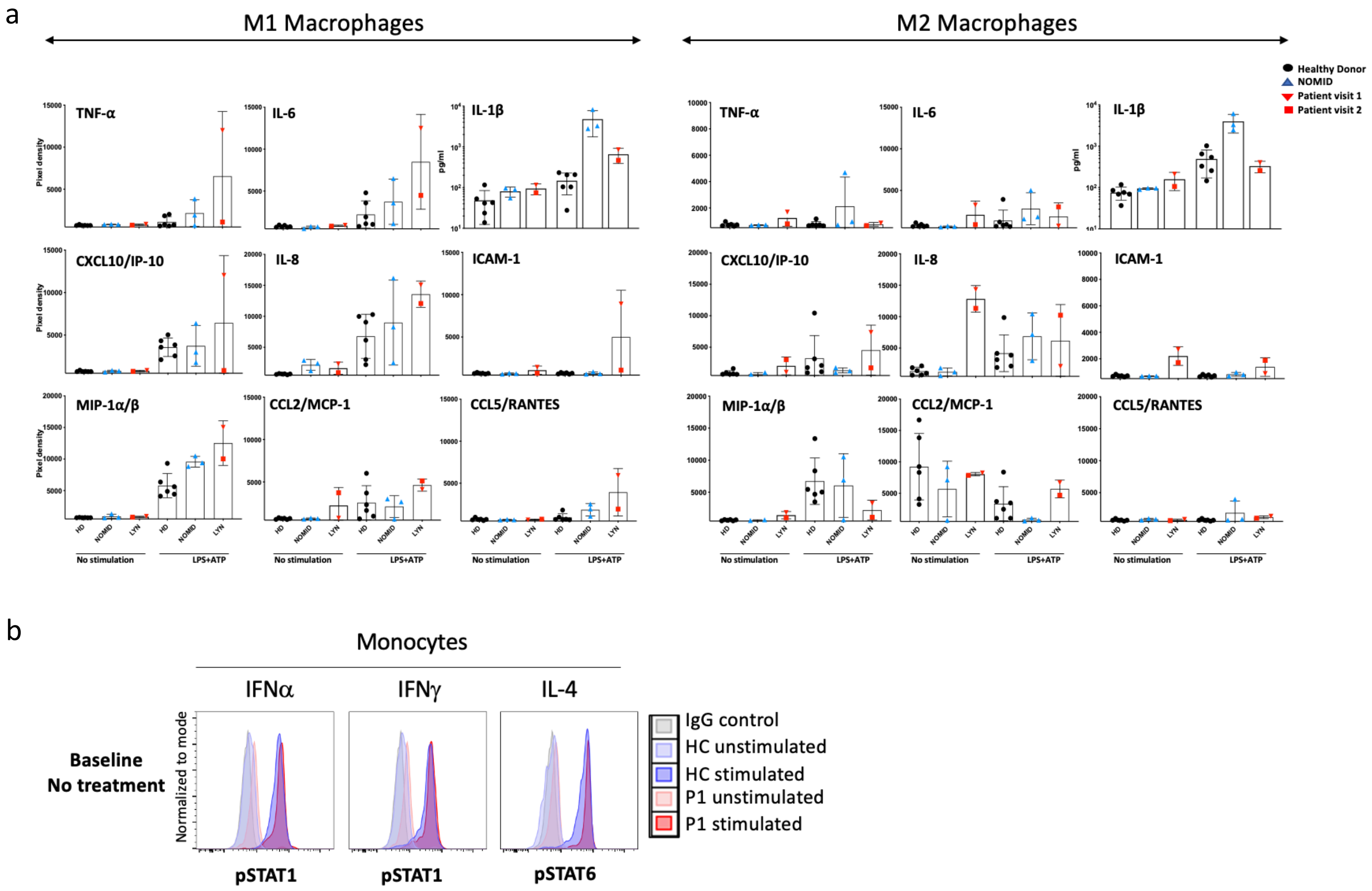


**a.** Stimulation of Patient 1's M1 and M2-like monocyte derived macrophages (MDM) with LPS resulted in higher IP-10, IL-6, TNF-$\alpha$ and mild IL-1$\beta$ overproduction predominantly in M1-like MDM when compared to healthy controls and to patients with the IL-1 mediated autoinflammatory disease NOMID. Patient 1 with the *LYN* GOF mutation had higher soluble ICAM-1 (sICAM-1) levels and shed sICAM-1 upon stimulation with LPS and ATP. Unstimulated M2-like MDM produced high levels of IL-8. **b.** Overlays of flow cytometry histograms of phospho-STATs (pSTAT1 and pSTAT6) signal in unstimulated (light color) and indicated cytokines-stimulated (dark color) peripheral blood mononuclear cells (PBMCs) from a healthy control and from the Patient 1. Gating was on monocytes by forward scatter (FSC) versus side scatter (SSC). Patient monocytes STAT1 and STAT6 phosphorylation was comparable to healthy control monocytes upon stimulation with IFN$\alpha$ and IFN$\gamma$ (pSTAT1) and IL-4 (pSTAT6).

#### Supplementary Figure 6. Effect of Various Treatments on Neutrophil Activation.

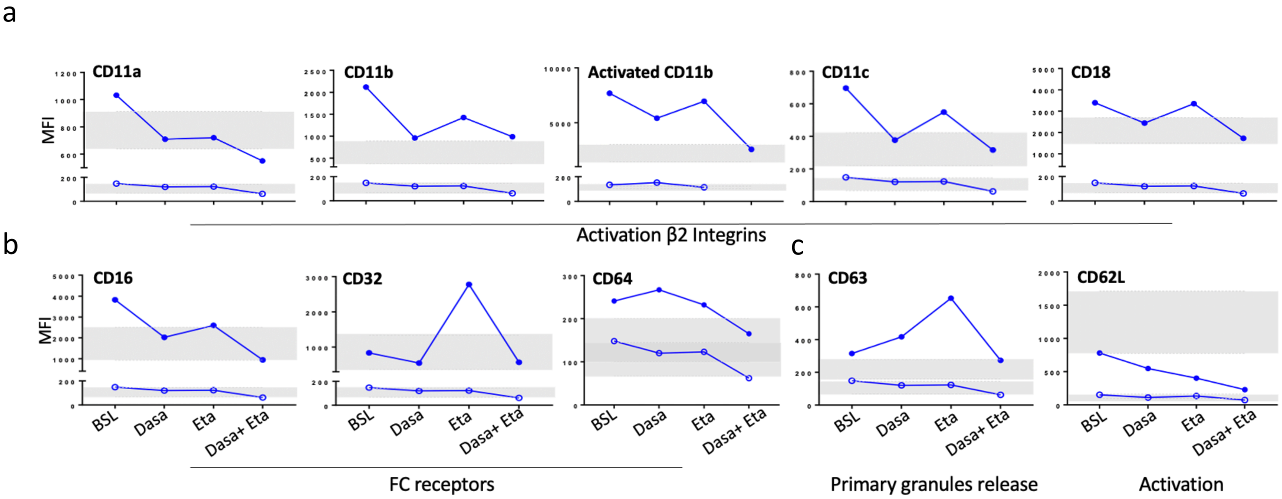


d e

| Endothelilal cells | | | | | Neutrophil | | | | |
|---|---|---|---|---|---|---|---|---|---|
| Soluble Receptor elevated at baseline | Alias | *Gene* | *Response to treatment* | **Effect of Etanercept vs Dasatinib** | Receptor | Alias | *Gene* | *Response to treatment* | **Effect of Etanercept vs Dasatinib** |
| **1. Tethering/Rolling** | | | | | | | | | |
| sE-selectin | sCD62E, SELE | *SELE* | (+/-) | *minimal improvement on Enbrel, dasatinib not done* | **L-selectin*** | CD62L, Leu 8 | *SELL* | low | remained low |
| sVCAM-1 | sCD106 | *VCAM1* | (-) | *no change* | | | | | |
| **3. Firm Adhesion/Crawling** | | | | | | | | | |
| sICAM-1 | CD54 | *ICAM1* | (+++) | *higher on no treatment, largest supression on etanercept plus dasatinib* | **αLβ2*** | **LFA-1,** CD11a/CD18 | *ITGAL, ITGB2* | (++) | improvement on etanercept, further improvement on etanercept and dasatinib |
| NA | | | NA | *NA* | **αMβ2*** | **Mac-1,** CD11b/CD18, Integrin Alpha-M, identical with CR3, the iC3b receptor, | *ITGAM, ITGB2* | (+++) | improvement on etanercept, normalization on etanercept and dasatinib |
| | | | | | **αxβ2*** | **CD11c/CD18,** Complement Component 3 Receptor 4 Subunit, alpha X, CR4 | *ITGAX, ITGB2* | (+++) | improvement on etanercept, normalization on etanercept and dasatinib |

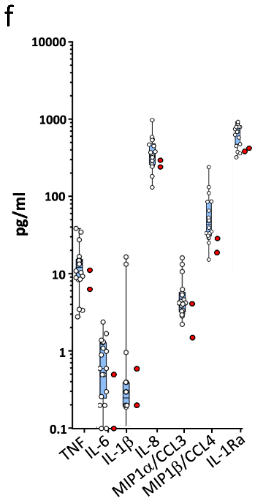


**a-e.** Patient 1's neutrophil surface markers were assessed by whole blood flow cytometry analysis at baseline (BSL), on dasatinib monotherapy (Dasa), etanercept monotherapy (Eta) and combination therapy of dasatinib and etanercept (Dasa+Eta). **a-c**. Open circles are isotype controls IgG1-FITC or IgG1-ACP, closed circles are specific antibodies. **a.** Neutrophils assessed at baseline expressed elevated levels of β2 integrins CD11a/LFA-1α, CD11b/Mac-1α, CD11c, and CD18/LFA-1β/Mac-1β. β2 integrin expression decreased with treatment especially with dasatinib as monotherapy or combined with etanercept. **b.** Neutrophils assessed at baseline had elevated levels of Fc gamma receptors I (FcγRI/CD64) and III (FcγRIII/CD16) and their expression normalized on dasatinib monotherapy or combined with etanercept. FcγRII/CD32 expression was normal at baseline and dasatinib monotherapy and increased on etanercept monotherapy, normal levels were achieved on combination therapy. **c.** Neutrophils had increased surface expression of CD63/LAMP3 indicating release of primary granules and neutrophil activation. Normal levels of CD63/LAMP3 were achieved with dasatinib and etanercept combination therapy (left panel). Constitutively low surface CD62L/Leu8/L-selectin indicates high soluble L-selectin that is shed from the neutrophil surface (right panel). **d.** Endothelial cell markers and effect of treatment on their expression is shown. **e.** Selected neutrophil activation markers depicted in **a-c** are described in detail and effect of treatment on their expression is shown. **f.** Cytokine production by unstimulated neutrophils from Patient 1 at two different time points (red symbols) is comparable to healthy controls (blue bar and white symbols) for the analytes tested. Patient was on dasatinib at the time of blood draw.

#### Supplementary Figure 7. Treatment Effect on Biliary Ductopenia and Liver Fibrosis in Patient 1.

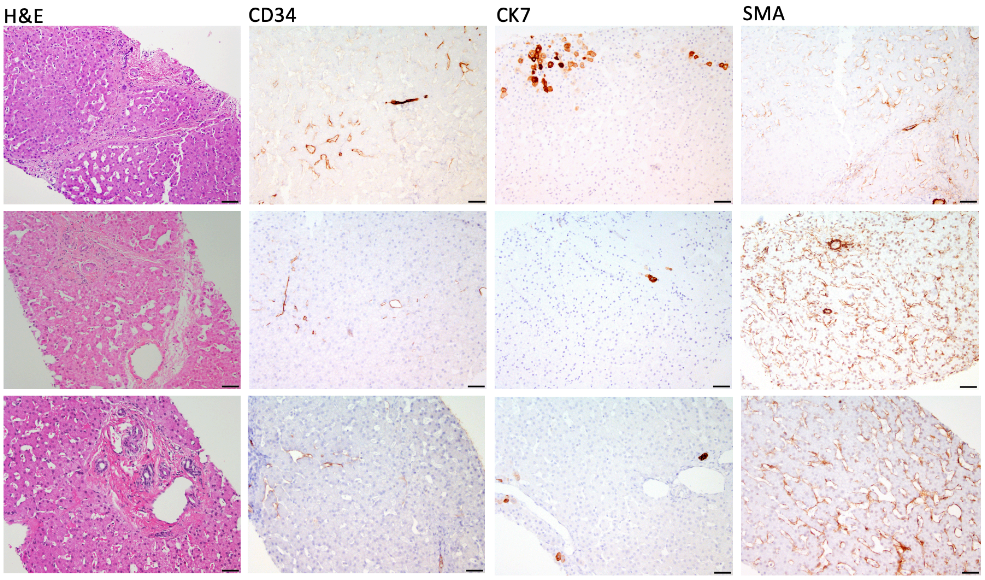


**Panels from top row to bottom**. Liver biopsy changes over time. Top row, biopsy after 6 months on glucocorticosteroids and IVIG (biopsy 2). The hepatic architecture is distorted by bridging fibrosis. CD34 and CK7 show focal abnormal staining of sinusoidal endothelial cells and hepatocytes respectively. SMA is expressed in only a minority of the stellate cells. Middle row, biopsy after 12 months on dasatinib treatment (biopsy 3). Bridging fibrosis remains, but the CD34 and CK7 stains have a more normal staining pattern. In contrast, SMA expression is now seen in most of the sinusoidal spaces. Bottom row, biopsy after 30 months on dasatinib monotherapy (biopsy 4). No bridging fibrosis was seen in the last biopsy and the portal areas have a normal configuration with normally sized portal veins. CD34 and CK7 are not significantly different from the prior biopsy. SMA staining of most of the stellate cells also persists. (All photos at 200x magnification, Scale bar = 50 microns).

#### Supplementary Figure 8. Lyn Mutations Increase Neutrophil Activation and Neutrophil Transendothelial Migration (TEM).

a

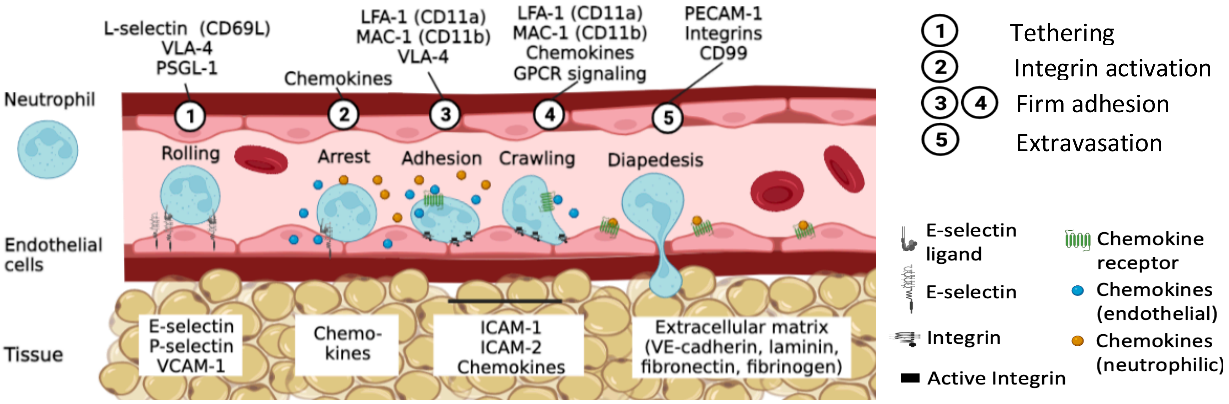


b c

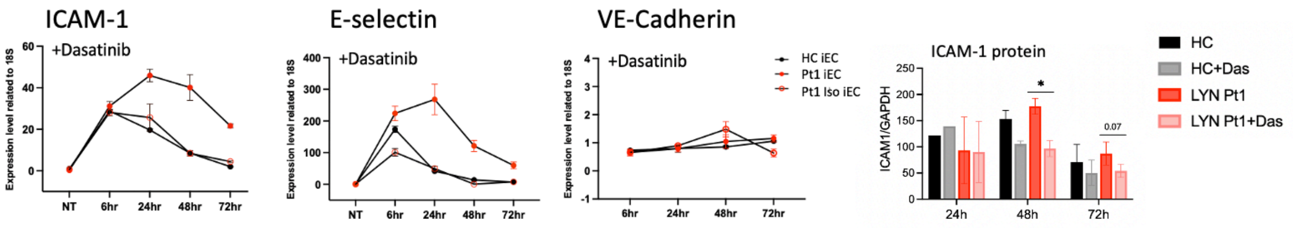


**a.** β2-integrins are required for the firm adhesion of neutrophils to the endothelial layer of blood vessels and for leukocyte extravasation into tissues.[7] This multistep process involves rolling (step 1), arrest (step 2) and firm adhesion (step 3) crawling (step 4), and extravasation/diapedesis (step 5).[8] Initially leukocyte engage with endothelial cells through selectins and VCAM-1 that allows rolling. Neutrophils also sense chemokines produced by the activated endothelium. In neutrophils, the binding to selectins and chemokine receptors activates β2-integrins into an intermediate affinity state, integrins bind to ICAM-1 on endothelial cells that mediates slow rolling, and into a high affinity state, which mediates leukocyte arrest.[9] This process ends with either transcellular or paracellular passage of the leukocyte across the endothelial monolayer. In the case of paracellular TEM, endothelial cell junctions are transiently disassembled to allow passage of leukocytes. **b.** mRNA expression of the endothelial cell markers ICAM-1, E-selectin, and VE-cadherin in Patient 1's iPS derived endothelial cells (Pt1 iEC), a corrected iEC clone (Pt1 Iso iEC) and a healthy control iEC (HC iEC) upon stimulation with IL-1β. Dasatinib does not reduce ICAM-1 transcription in IL-1β stimulated wildtype (HC and Pt1 Iso iEC) and mutant (Pt1 iEC) iECs. **c.** ICAM-1 protein expression on IL-1β stimulated patient and control iECs was quantified by Western blot and significantly decreased with addition of dasatinib that was most pronounced at 48 hrs.

#### Supplementary Figure 9. Assessment of Vascular Permeability by VE-Cadherin Staining and by Transendothelial Resistance by Electric Cell-Substrate Impedance Sensing (ECIS).

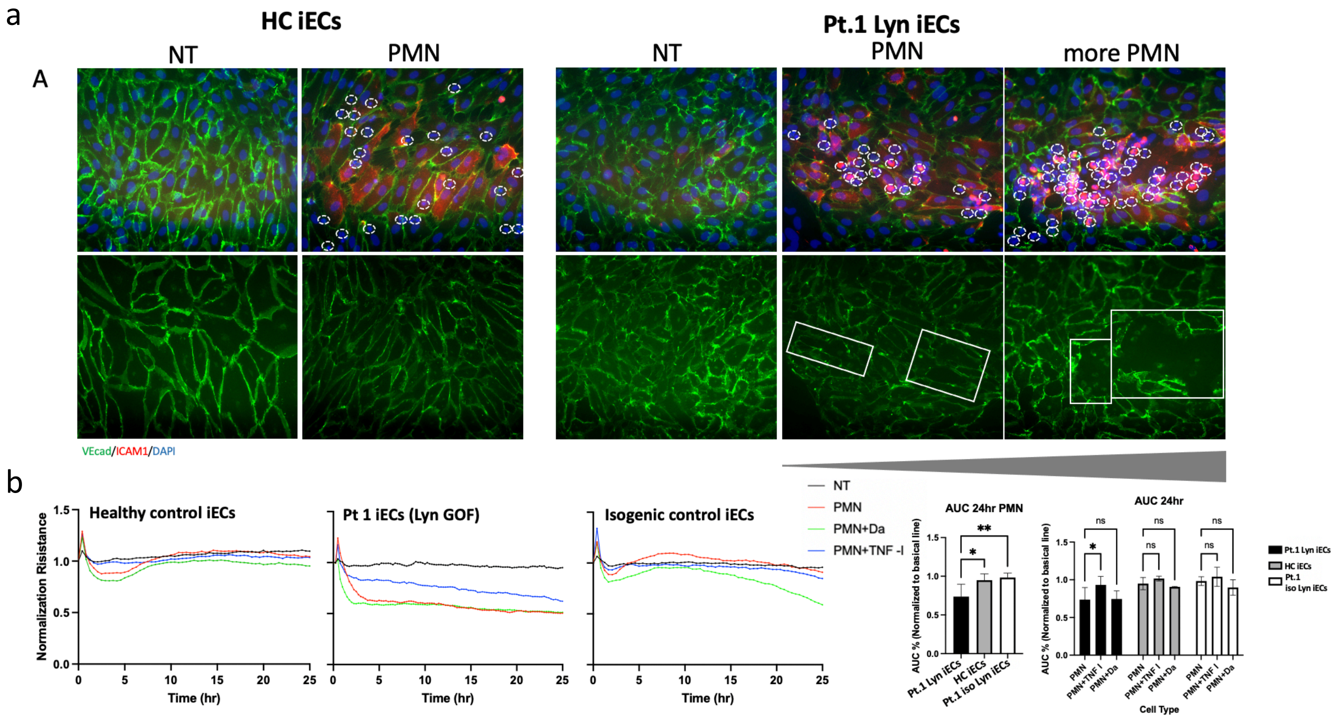


**a.** Co-culture of neutrophils with iECs leads to decreased VE-cadherin staining (green) in Patient 1 iECs (white boxes) compared to healthy control iECs which was more pronounced in areas of increased neutrophil adhesion. The internalization of VE-cadherin into subcellular compartments and subsequent recycling or degradation and the concomitant disruptions of cell–cell contacts have been linked to increased vascular permeability[10] which was reflected in reduced transendothelial electrical resistance (TEER).[11] Gray triangle indicates increasing number of adherent neutrophils. **b.** We thus conducted real time measurement of transendothelial electrical resistance (TEER) across the endothelial monolayer using Electric Cell-substrate Impedance Sensing (ECIS). ECIS Zθ technology is used to assess and quantify endothelial permeability. ECIS data presentation of baseline absolute resistance (ohm, Ω) in iPS cell derived endothelial cell (iEC) from Patient 1 (LYN GOF), a healthy control (HC), and an isogenic control originating from LYN Patient 1 (Pt.1 iso Lyn) were assessed upon co-culture with healthy control neutrophils (polymorphonuclear cells, PMN) without and with treatment with a TNF inhibitor (TNF-I) or with dasatinib (Da). All absolute values were normalized to the last reading before the additional treatment. The changes and influence of resistance were analyzed using area under curve (AUC). Left panels are the representative profile of endothelial barrier resistance. Black line: iEC alone; red line: iEC PMN added; blue line: PMN plus TNF inhibitor; green line: PMN plus dasatinib. Right panels show changes of resistance in iEC (Mean ± SD, n=5). Addition of neutrophils to iEC culture leads to significantly decreased transendothelial resistance in Pt. 1 iECs compared to HC iECs ($p<0.05$) and Pt.1 iso iECs ($p<0.01$). Addition of TNF inhibitor but not dasatinib to the iEC and neutrophil co-culture significantly increased resistance in Pt.1 iECs ($p<0.05$).

### III. Supplementary Tables

#### Supplementary Table 1. Features of the *LYN* Mutations Detected in Patients 1,2,3.

| **Gene** | ***LYN*** | | |
|---|---|---|---|
| **pLI** | 1.00 | | |
| **Transcript** | NM_002350.4, ENST00000519728.6 | | |
| **Patient** | Pt.3 | Pt.2 | Pt.1 |
| **Chr coordinates (GRCh37)** | Chr8:56922649 C>T | Chr8:56922653 A>T | Chr8:56922654 C>G |
| **HGVSc** | c.1519C>T | c.1523A>T | c.1524C>G |
| **HGVSp** | p.Q507* | p.Y508F | p.Y508* |
| **Inheritance mode** | De novo dominant | De novo dominant | De novo dominant |
| **rsID** | NA | NA | NA |
| **gnomAD AF** | 0 | 0 | 0 |
| **GERP score** | 5.95 | 5.95 | 5.95 |
| **CADD phred** | 40 | 28 | 36 |
| **MetaSVM_prediction** | NA | Deleterious | NA |
| **MutationTaster_prediction** | Disease causing | Disease causing | Disease causing |
| **PROVEAN_prediction** | NA | Deleterious | NA |
| **Polyphen2_HDIV_prediction** | NA | Prob_damaging | NA |
| **SIFT_prediction** | NA | Deleterious | NA |
| **BLOSUM62** | NA | 3 | NA |
| **MSC-CADD damage prediction** | High impact | High impact | High impact |
| **Present in COSMIC** | No | No | No |

pLI: probability of loss-of-function intolerance. *LYN* is intolerant to haploinsufficiency as indicated by a "probability of being loss-of-function intolerant" [pLI] score of 1.0, on a range of 0.0 to 1.0, with a higher score indicating a greater degree of intolerance for loss-of-function variants in healthy persons.
HGVS: Human Genome Variation Society.
gnomAD_AF: Genome Aggregation Database (gnomAD) v.2.1.1 minor allele frequency for all populations
GERP: Genomic Evolutionary Rate Profiling. Score threshold of 2 or greater indicates truly constrained sites.
CADD_phred: CADD Whole-genome Combined Annotation-Dependent Depletion (CADD) scores are based on conservation metrics, functional genomic data, transcript information, and protein-level scores (e.g. SIFT, and PolyPhen). Higher CADD_raw and CADD_phred scores indicate that a variant is more likely to be deleterious, CADD_phred scores >=10 predict that a variant is amongst the 10% most deleterious of all possible substitutions, CADD_phred scores >=20 predict that a variant is amongst the 1% most deleterious of all possible substitutions.[12]
MetaSVM: Radial kernel support vector machine network based on nine prediction scores and allele frequencies in 1000G.[13]
PROVEAN (Protein Variation Effect Analyzer) is a software tool that predicts whether an amino acid substitution or indel has an impact on the biological function of a protein.[14]
PolyPhen2_HDIV: Polymorphism Phenotyping v2.[15]
SIFT: Sorting Intolerant From Tolerant.[16]
BLOSUM62: BLOcks of Amino Acid SUbstitution Matrix, a positive BLOSUM62 score means that the amino acid substitution is conservative, the two amino acids are biochemically similar
MSC-CADD: Mutation Significance Cutoff-Combined Annotation Dependent Depletion
COSMIC: Catalogue of Somatic Mutations in Cancer

#### Supplementary Table 2. Broad Assessment of Neutrophil Function.

| *Functional tests were performed while patient was on either dasatinib or etanercept treatment** | *Result* |
|---|---|
| **1. ROS production by luminol-enhanced chemiluminescence (3 time points)** | within normal limits |
| **2. PMN adherence to plastic coated with extracellular matrix proteins** | within normal limits |
| **3. Adherent superoxide production (3 time points)** | within normal limits |
| **4. Neutrophil migration (chemotaxis by EZ-TAXIScan in response to fMLF)** | not different from control neutrophils |
| **5. Broad neutrophil stimulation assaying IL-8, IL-1β, IL1RA, IL-6, MIP1α, MIP1β, and TNF production in response to a broad set of stimuli**** | within normal limits |
| **6. Phagocytosis** | not affected |
| **7. Degranulation (plasma level of granule enzymes)***** | constitutive degranulation higher than in controls |

* The assays are limited by the fact that the patient has been on treatment at the time of the evaluation, however treatment with either etanercept or dasatinib did not impact the stimulation assay differently
** Stimulation assays with TLR agonists, microbes, other DAMPs (see supplementary methods)
*** Neutrophil granule enzymes measured: MMP-9, lipocalin/NGAL, lactoferrin, MPO, RANTES

#### Supplementary Table 3. Summary of Liver Biopsies with Progression of Liver Disease Prior to Dasatinib Treatment.

| **Biopsy number** | **Ishak Fibrosis Stage** | **Duct count** | **Treatment** |
|---|---|---|---|
| **1** | Periportal fibrosis stage 1 | 15 ducts/26 portal areas (*57%*) | No treatment |
| **2** | Bridging fibrosis stage 3 | 3 ducts/6 portal areas (*50%*) | 6 mos on GC + IVIG |
| **3** | Bridging fibrosis stage 3 | 14 ducts/18 portal areas (*78%*) | 12 mos on dasatinib |
| **4** | Periportal fibrosis stage 1 | 12 ducts/13 portal areas (*92%*) | 30 mos on dasatinib |

mos: months; GC: glucocorticosteroids; IVIG: intravenous immunoglobulin

**Supplementary Table 4. Effects of Various Treatments on Co-culture of Healthy Control or Patient 1 Neutrophils with Patient 1 induced pluripotent stem (iPS) cell-derived endothelial cells (iECs).**

| Intervention | Inflammation | Adhesion* | TEER* reduced VE-cadherin expression | TEM* |
|---|---|---|---|---|
| Neutrophil co-culture | Pt neutrophils induce IL-6 | Increased in Lyn iECs | Necessary to decrease TEER in Lyn iECs | Increased in Lyn iECs |
| + Dasatinib | **Blocks IL-6 production** | **Blocks adhesion** | No effect | Improves TEM by ~50% |
| + TNF inhibition | Minimal effect | Minimal effect | **Improves TEER in coculture in Lyn iECs** | Improves TEM by ~50% |
| + Dasatinib and TNFi | No synergism | No synergism | No synergism | No synergism |

*These experiments were only performed with healthy control neutrophils. Due to clumping of the patient neutrophils upon isolation these experiments could not be performed.
iEC: induced pluripotent stem (iPS) cell-derived endothelial cells; TEER: transendothelial electrical resistance; TEM: transendothelial migration

### IV. Supplementary Results

#### Evaluation of Autoimmune Dysregulation

Given the literature on the role of Lyn kinase in driving autoimmunity, we evaluated the B cell compartment clinically and by performing B cell stimulation assays in Patient 1 (Supplementary Figures S10 and S11). Patient 1 developed autoantibodies after splenectomy. He had positive antinuclear (ANA), and low titer anti-Sm, anti-SSA, anticardiolipin IgG, and anti-mitochondrial antibodies, lupus anticoagulant, and rheumatoid factor. After 7 months on dasatinib, except for ANA, all autoantibodies were negative, and ANA turned negative after 3.5 years on dasatinib therapy and turned positive during the 11 months the patient was on etanercept monotherapy.
Patient 2 never had positive autoantibodies on clinical tests and patient 3 had a borderline anti-PR3 antibody that has not been repeated.
Spleen histology was assessed in Patient 1 (Supplementary Figure S12), who was splenectomized, and showed a disrupted spleen architecture. Consistent with the $Lyn^{up/up}$ or $Lyn^{+/up}$ mouse,[17] Patient 1's spleen has poorly formed lymphoid follicles and lacks germinal centers, evidenced by the abnormally distributed CD20 and CD21, and negative BCL6 immunostainings, respectively. Naïve B cells (IgD+) are also abnormally distributed in the spleen lymphoid follicles, similarly to the $Lyn^{up/up}$ or $Lyn^{+/up}$ mice, which also display reduced marginal zone B cell numbers.[17] Whether the disorganization of the spleen architecture in Patient 1 is caused by a Lyn kinase associated migration or trafficking defect of B cells and possibly follicular dendritic cells, which are essential for spleen germinal center formation, needs to be further assessed.

**Supplementary Figure i: Assessment of B Cell Function in Bone Marrow and Peripheral Compartment.**

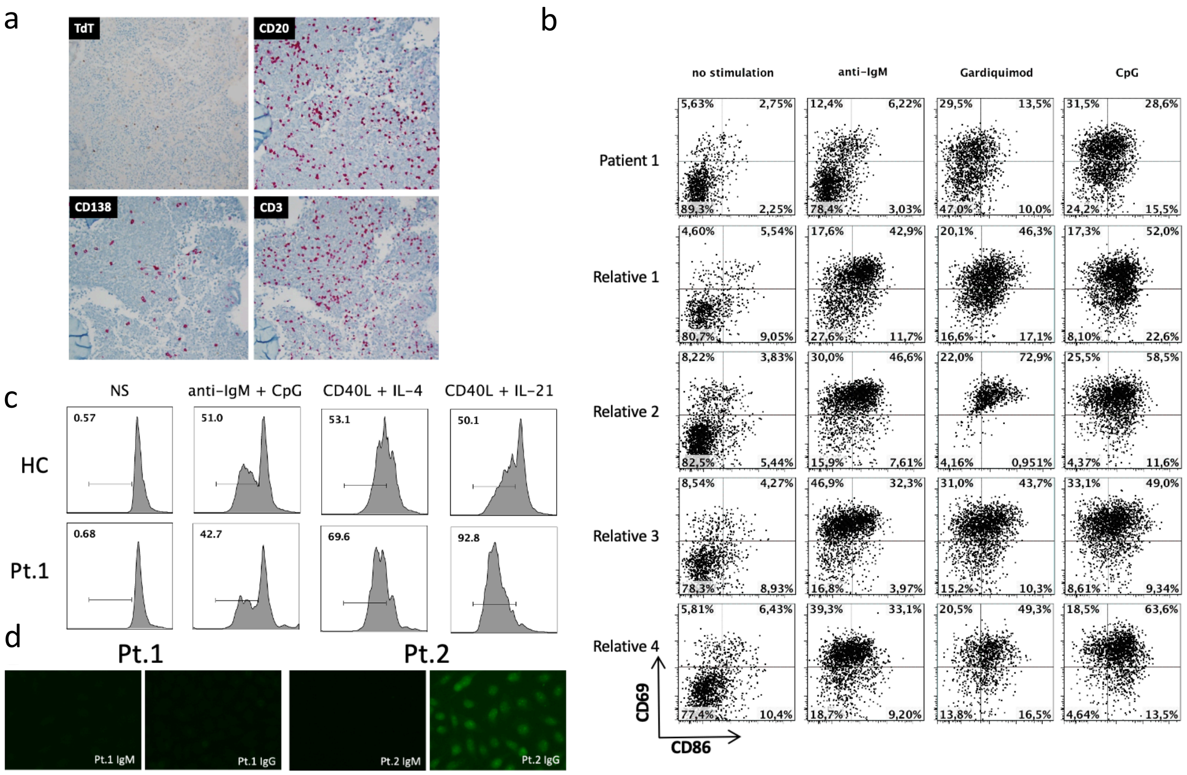


**a.** Sections of the bone marrow core biopsy have 100% cellularity. TdT (hematogones) demonstrates few B-cell precursors consistent with the high doses of prednisone 1mg/kg/day the patient was receiving at the time of the biopsy. CD20 stains scattered mature B-cells (CD79a showed similar pattern). CD138 stains occasional scattered plasma cells. CD3 stains scattered T-cells which are similar in numbers to B-cells which is reduced compared to age matched controls. **b**. B-cells isolated from peripheral blood were stimulated with anti-IgM (B cell receptor) or the TLR agonists, Gardiquimod (TLR7) or CpG (TLR9) respectively. The patients' BCR function are reduced in response to anti-IgM, Gardiquimod or CpG stimulation. These findings are consistent with a defective central B cell tolerance (Eric Meffre personal communication). **c.** B cells from patient 1 proliferate more when stimulated with cytokines especially IL-21, which may account in part for the autoantibody secretion in this patient. **d.** ANA testing by Hep2 cell staining shows serum from Patient 1 to be negative for ANA and for Patient 2 to be positive for ANA IgG antibodies. HC=healthy control, Pt.1 = Patient 1, Pt.2 = Patient 2, suggesting transient ANA positivity.

**Supplementary Figure ii: Evaluation of Treg Cell Function in Patient 1 Compared to a Healthy Control.**

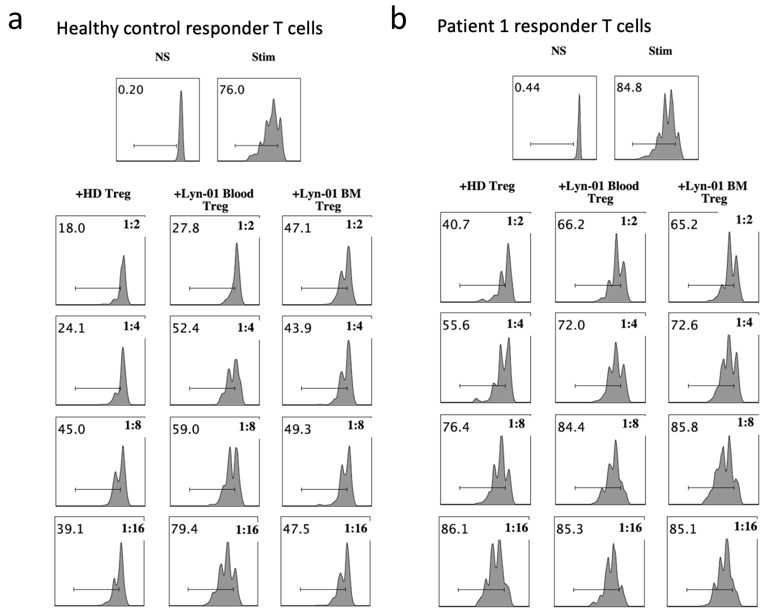


Assessment of *in vitro* Treg suppressive function. **a**. Tregs isolated from both blood and BM of patient 1 had suboptimum abilities to inhibit the proliferation of CFSE-labeled CD3$^{+}$CD4$^{+}$CD25$^{-}$ autologous and heterologous Tresponder (Tresp) cells compared to Tregs isolated from the blood of a healthy donor control (HD). **b.** In addition, Tresp from Patient 1 were refractory to suppression by HD Tregs.

##### Supplementary Figure iii: Spleen Histology of Patient 1 with Splenectomy

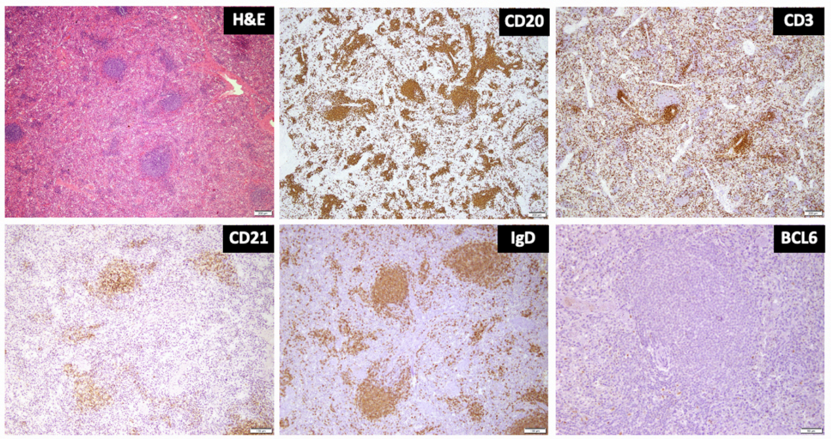


**Upper left panel**: H&E staining of the spleen shows that the white pulp is not well expanded and small lymphoid aggregates are present in the red pulp, **Upper center panel:** CD20 staining depicts that the aggregates are mostly composed of B cells. **Upper right panel**: CD3 staining shows normal peri-arteriolar sheath distribution of T cells. **Lower left panel:** CD21 staining shows some follicular dendritic cells in poorly formed follicles, not well organized as typically seen in a secondary follicle with a well-formed germinal center. **Lower center panel:** IgD staining highlights abnormally distributed naïve B cells indicating that the patient does not form secondary follicles. **Lower center panel:** The lack of well-formed germinal centers is confirmed by the absence of BCL6 staining in the primary follicles.
